# Panorama of Chromosomal Instability in Lung Cancer

**DOI:** 10.1101/2025.09.09.25335109

**Authors:** Yang Yang, Xiaoming Zhong, William Phillips, Wei Zhao, Phuc H. Hoang, Christopher Wirth, Soo-Ryum Yang, Charles Leduc, Marina K. Baine, William D. Travis, Lynette M. Sholl, Philippe Joubert, Robert Homer, Jian Sang, Azhar Khandekar, John P. McElderry, Thi-Van-Trinh Tran, Caleb Hartman, Mona Miraftab, Monjoy Saha, Olivia W. Lee, Sunandini Sharma, Kristine M. Jones, Bin Zhu, Marcos Díaz-Gay, Eric S. Edell, Jacobo Martínez Santamaría, Matthew B. Schabath, Sai S. Yendamuri, Marta Manczuk, Jolanta Lissowska, Beata Świątkowska, Anush Mukeria, Oxana Shangina, David Zaridze, Ivana Holcatova, Vladimir Janout, Dana Mates, Simona Ognjanovic, Milan Savic, Milica Kontic, Yohan Bossé, Bonnie E. Gould Rothberg, David C. Christiani, Valerie Gaborieau, Paul Brennan, Geoffrey Liu, Paul Hofman, Maria Pik Wong, Kin Chung Leung, Chih-Yi Chen, Chao Agnes Hsiung, Angela C. Pesatori, Dario Consonni, Nathaniel Rothman, Qing Lan, Martin A. Nowak, David C. Wedge, Ludmil B. Alexandrov, Stephen J. Chanock, Jianxin Shi, Tongwu Zhang, Lixing Yang, Maria Teresa Landi

## Abstract

Lung cancer is a highly heterogeneous disease primarily driven by tobacco smoking. About 20% of lung cancers occur among patients who have never smoked (LCINS) with differences in patient ancestry, sex, tumor histology, and clinical features. Our understanding of chromosomal instability in lung cancer, especially LCINS, is still limited. Here, we perform a comprehensive study of 182,429 somatic structural variations (SVs) detected in 1,209 whole-genome sequenced lung cancers, of which 864 LCINS. SVs are more abundant in tumors from patients who have smoked (LCSS); however, they are more complex and play more important roles in tumorigenesis in LCINS. *EGFR* mutations and *KRAS* mutations profoundly and independently shape the SV landscape. *EGFR*-mutant tumors have higher SV burden and more cancer-driving SVs. In contrast, *KRAS* mutations are associated with lower SV burden and less driver SVs. We decompose 16 SV signatures for both complex and simple SVs that likely represent divergent molecular mechanisms. The SV breakpoints have distinct distributions across the genome depending on the signatures due to mutagenic mechanisms and positive selection. Many established cancer-driving genes are recurrently rearranged by multiple SV signatures suggesting functional convergence of these genome instability mechanisms.

## Introduction

Lung cancer is the leading cause of cancer death worldwide in both men and women^1^. Although most lung cancer diagnoses are attributed to tobacco smoking, approximately 15% of male patients and 50% of female patients have never smoked^2^ (defined as having smoked fewer than 100 cigarettes in their lifetime). Large geographic variations in incidence rates of lung cancers in patients who have never smoked (LCINS) have been documented^3^. LCINS are biologically and clinically distinct in many aspects. For example, all major histological types of lung cancers (small cell lung cancer, squamous cell carcinoma, adenocarcinoma and large cell carcinoma) are associated with smoking; however, LCINS are predominantly adenocarcinomas^4,5^. Genetically, *EGFR* mutations are enriched in LCINS^6^, while *KRAS* and *TP53* mutations are more common in tumors from patients who have smoked^7,8^ (LCSS). Clinically, patients who have never smoked tend to have better survival^9,10^ and superior response to EGFR inhibitors^11,12^, but less durable response to immune checkpoint blockade^13,14^.

Genome instability is a well-established hallmark of cancer^15,16^ and somatic structural variations (SVs) are abundant in adult solid tumors^17^. SVs refer to alterations in the DNA sequence that involve large segments of a genome. They include deletions (Del), tandem duplications (TD), inversions (Inv), translocations (Tra) and other complex events. Somatic SVs are of clinical significance in LCINS due to the high frequency of oncogenic fusions caused by SVs^18–20^. Our knowledge of the underlying causes of somatic SVs and their contributions to genome instability is rather limited. Different mutational processes leave distinct footprints on genomic DNA which can be mathematically decomposed into mutational signatures. Somatic SV and somatic copy number alteration (SCNA) signatures have been deconvoluted in multiple pan-cancer studies^21,22^ without distinguishing complex and simple SVs. Complex SVs consist of multiple SV breakpoints produced all together as one-time events, such as chromothripsis^23^ and chromoplexy^24^. They arise through distinct molecular mechanisms. Some are clustered on one or a few chromosomes whereas others are scattered. Chromosomes in micronuclei - formed through lagging chromosomes during mitosis - and chromatin bridges -formed through dicentric chromosomes - can shatter and form chromothripsis^25,26^. Once chromosomes are shattered, some DNA fragments can be ligated into a circular form known as circular extrachromosomal DNA (ecDNA)^25^, which is often highly amplified in cancer^27^. ecDNA can sometimes integrate into linear chromosomes and form homogeneous staining regions (HSRs)^28^. Furthermore, chromoplexy was proposed to form through reciprocal translocations involving multiple partners simultaneously and the breakpoints are scattered across multiple chromosomes^24^. We call ‘clustered complex SVs’ the complex SVs that have breakpoints enriched in certain chromosomes more than expected by chance, such as chromothripsis. The complex SVs without breakpoint enrichment, for example, chromoplexy, are considered ‘non-clustered complex SVs’.

In this study, we performed a detailed analysis on somatic SVs in 1,209 whole-genome sequenced lung cancers, including 864 LCINS^29^. We separately characterized clustered complex SVs, non-clustered complex SVs, and simple SVs to more effectively elucidate the molecular mechanisms underlying SV formation and to better define their respective contributions to tumorigenesis.

## Results

### Study cohort and somatic SVs

To comprehensively study the somatic SV landscape in lung cancers, we collected a total of 1,209 cases, including 1,056 cases from the Sherlock-*Lung* Project (primarily LCINS)^29–31^ and 153 cases from published studies (mainly LCSS)^32–37^ (**Supplementary Table S1**). Patients were recruited from 30 different locations across four continents. Among these, 811 and 359 patients had European (EUR) and East Asian (EAS) genetic ancestries, respectively (**Fig. 1a**). The remaining 39 patients had South American and African genetic ancestries. Patients of European descent were almost exclusively recruited from Europe, Russia, USA and Canada, while patients of East Asian descent were recruited mainly from Hong Kong, Taiwan, South Korea and Canada (**Supplementary Table S1**). The dataset included 864 LCINS^29^ and 344 LCSS^31^. One subject had unknown smoking status. The majority (n = 1,032) of the tumors were adenocarcinomas, followed by 71 squamous cell carcinomas, 60 carcinoid tumors, 11 adenosquamous carcinomas and 35 of other histologies (**Fig. 1a**). Both tumor and matched normal samples were whole genome sequenced at an average depth of 80x and 40x coverage, respectively.

**Figure 1.**
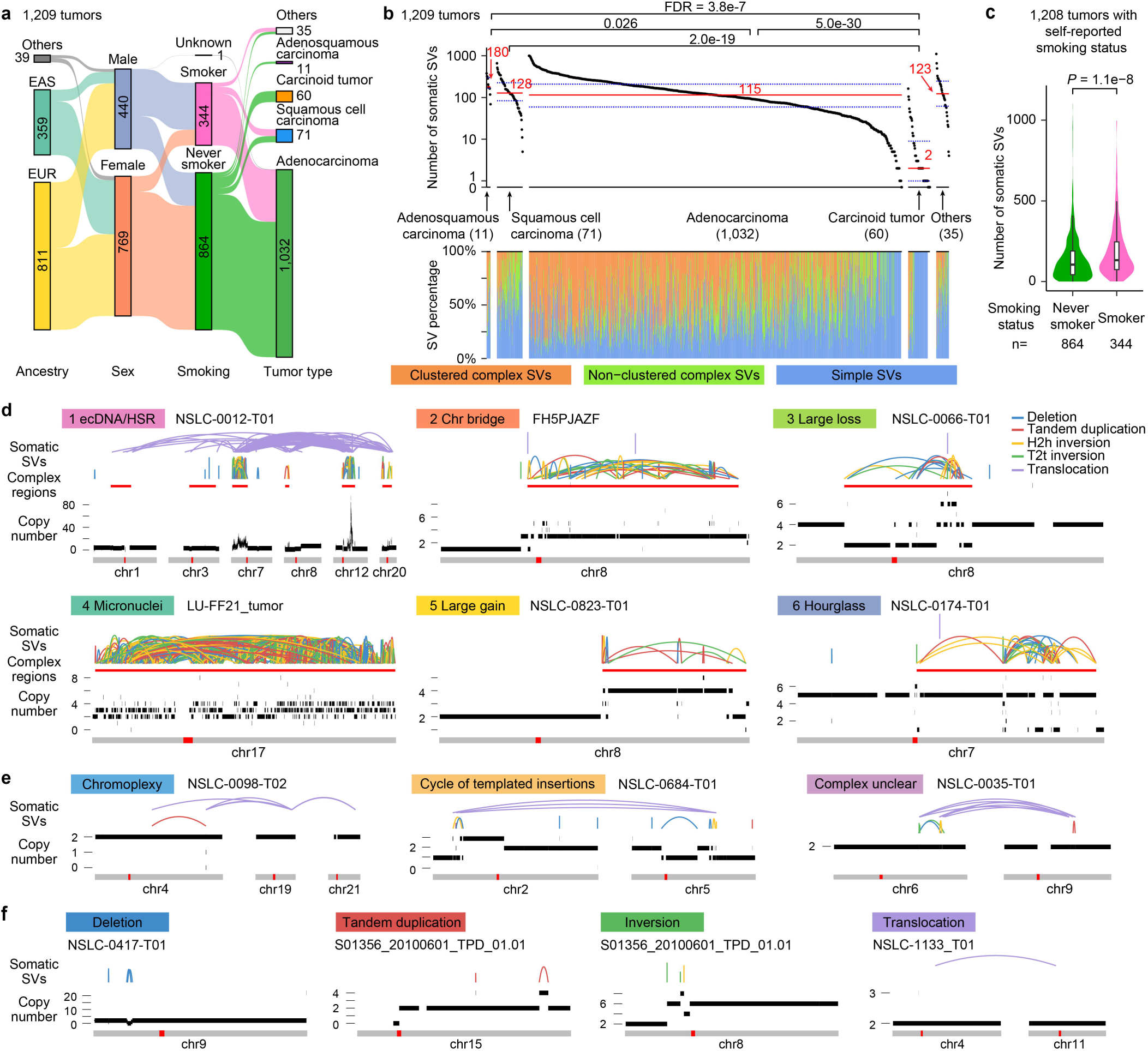
Landscape of somatic SVs in 1,209 whole-genome sequenced lung cancers. **a**, Sankey diagram illustrates the relationships among ancestry, sex, self-reported smoking status and tumor histology type in our cohort of 1,209 lung cancers. The vertical bars represent four categories (ancestry, sex, smoking and tumor type). The height of each bar is proportional to the number of patients/tumors in each category. The connecting bands represent the flow of patients/tumors from one category to another. The thickness of each link is proportional to the number of patients/tumors transitioning between the connected nodes. **b**, SV burden and composition across tumor types. The black dots on the top panel depict the number of somatic SVs detected in each tumor grouped by tumor type. Red lines indicate the median numbers of SVs, while blue dashed lines indicate the interquartile range (IQR) boundaries (Q1 and Q3). Two-sided Wilcoxon rank-sum test with False Discovery Rates (FDR) correction is used to compare SV burdens across tumor types (excluding “Others”), with only FDRs < 0.05 shown. The lower panel shows the composition of SVs, including clustered complex SVs, non-clustered complex SVs and simple SVs. **c**, SV burden in LCINS and LCSS. Box plots within violins indicate the interquartile ranges and medians. A two-sided Wilcoxon rank-sum test is used to calculate the *P* value. **d, e** and **f**, Examples of clustered complex SVs (**d**), non-clustered complex SVs (**e**) and simple SVs (**f**). Colored arcs represent SVs of different types. Red bars beneath the arcs in (**d**) mark the regions of clustered complex SVs. The copy number profiles are shown as black bars above the chromosome models. Centromere positions are highlighted as red bars within the grey chromosome ideograms. Sample IDs are displayed next to the corresponding SV signatures, and specific SV signatures/types are labeled for each example (e.g., 1 ecDNA/HSR, chromoplexy, deletion, etc.).

A total of 182,429 somatic SVs were detected in 1,209 tumors. The number of SVs (ranging from 0 to 1,102) varied significantly across histology with medians of 180, 128 and 115 for adenosquamous carcinomas, squamous carcinomas and adenocarcinomas, respectively (**Fig. 1b**). Carcinoid tumors had the fewest SVs (median of 2). In addition, driver fusions were detected in 95 tumors (7.9%) (**Supplementary Table S2**), highlighting the impact of SVs in lung cancer. LCSS had significantly higher SV counts than LCINS (*P*=1.1 × 10^-8^, **Fig. 1c**). To gain further insights into smoking status, we assessed single-base substitution signature 4 (SBS4), which is known to be mostly driven by tobacco smoking^38^. Most tumors from self-reported never smokers did not carry SBS4 mutations and those from smokers carried more than 100 mutations of SBS4 as expected (**Extended Data Fig. S1a**). However, there were 55 LCINS (6.4%) with more than 100 mutations of SBS4, whereas 39 LCSS (11.3%) lacked SBS4. If mis-annotation or inaccurate self-reporting of smoking status were the primary causes of the inconsistency between smoking status and SBS4, the mutation spectrum would be expected to be similar in tumors defined by SBS4 but different in self-reported smoking status. Intriguingly, this was not the case. The primary type of *EGFR* mutation in LCINS with SBS4 was L858R, which was barely observed in LCSS with SBS4^29^ (*P*=3.7 × 10^-4^, **Extended Data Fig. 1b**). Therefore, it is likely that at least a subset of patients who have never smoked were not misannotated, and the SBS4 mutations may be derived from passive smoking, air pollution, or other exposures^29^. Furthermore, driver fusions were found in 28.2% of LCSS without SBS4, which was much higher than that in LCINS without SBS4 (*P*=1.3 × 10^-3^, 9.6%) (**Extended Data Fig. 1b**). LCSS without SBS4 may have originated before patients began smoking. It is also possible that these tumors originated from cells not influenced by smoking, because it has been reported that a small fraction of pathologically normal bronchial epithelial cells in individuals with smoking history have mutation burdens similar to LCINS with very few SBS4 mutations^39^. Hence, tumors with SBS4 status different from self-reported smoking status may represent unique subtypes. In the remainder of this manuscript, SV signatures are decomposed for all tumors, but only 1,114 tumors with consistent self-reported smoking status and SBS4 were used to study the impacts of smoking on the repertoire of SVs. Furthermore, only EUR and EAS patients are used when genetic ancestry is studied as a factor.

### Complex SV events

Out of 182,429 individual somatic SVs, 91,776 (50.3%) and 25,652 (14.1%) belonged to clustered (**Fig. 1d**) and non-clustered (**Fig. 1e**) complex SVs, respectively. The remaining 65,001 (35.6%) SVs were classified as simple SVs (**Fig. 1f**). A total of 1,457 clustered complex SV events from 928 (76.8%) tumors and 4,816 non-clustered complex SV events from 1,053 (86.6%) tumors were detected (**Fig. 2a** and **Supplementary Tables S3, S4**). We then used Starfish^40^ and ClusterSV^22^ to identify clustered (**Fig. 1d**) and non-clustered (**Fig. 1e**) complex SV signatures, respectively, which likely reflect their molecular mechanisms of formation. The most abundant signature of clustered complex SVs was Signature 2 chromatin bridge detected in 310 (25.6%) tumors (**Fig. 2a**). Signature 1 ecDNA/HSR, Signature 5 large gain, Signature 4 micronuclei, Signature 6 hourglass and Signature 3 large loss were found in 278 (23.0%), 222 (18.4%), 209 (17.3%), 170 (14.1%) and 142 (11.7%) tumors, respectively (**Fig. 2a**). We note that “ecDNA/HSR” here refers to a signature of clustered complex SVs with both circular and linear forms. The simple form of ecDNA, one or a few DNA fragments ligated in a circle, was not captured by this signature and is analyzed in detail in a companion paper^41^. Regarding non-clustered complex SVs, 487 (40.3%) and 587 (48.6%) tumors carried chromoplexy and cycle of templated insertions (possibly formed through a template-switching mechanism^42^), respectively (**Fig. 2a**). Non-clustered complex SVs not classified as chromoplexy or cycle of templated insertions were assigned as “complex unclear” which may form through multiple unknown mechanisms. Both clustered and non-clustered SVs were more complex in LCINS than in LCSS (*P*=1.2 × 10^-4^ and 0.035, **Fig. 2b**). Among LCINS, clustered SVs were more complex in tumors from EAS patients than those from EUR patients (*P*=0.012, **Fig. 2b**).

**Figure 2.**
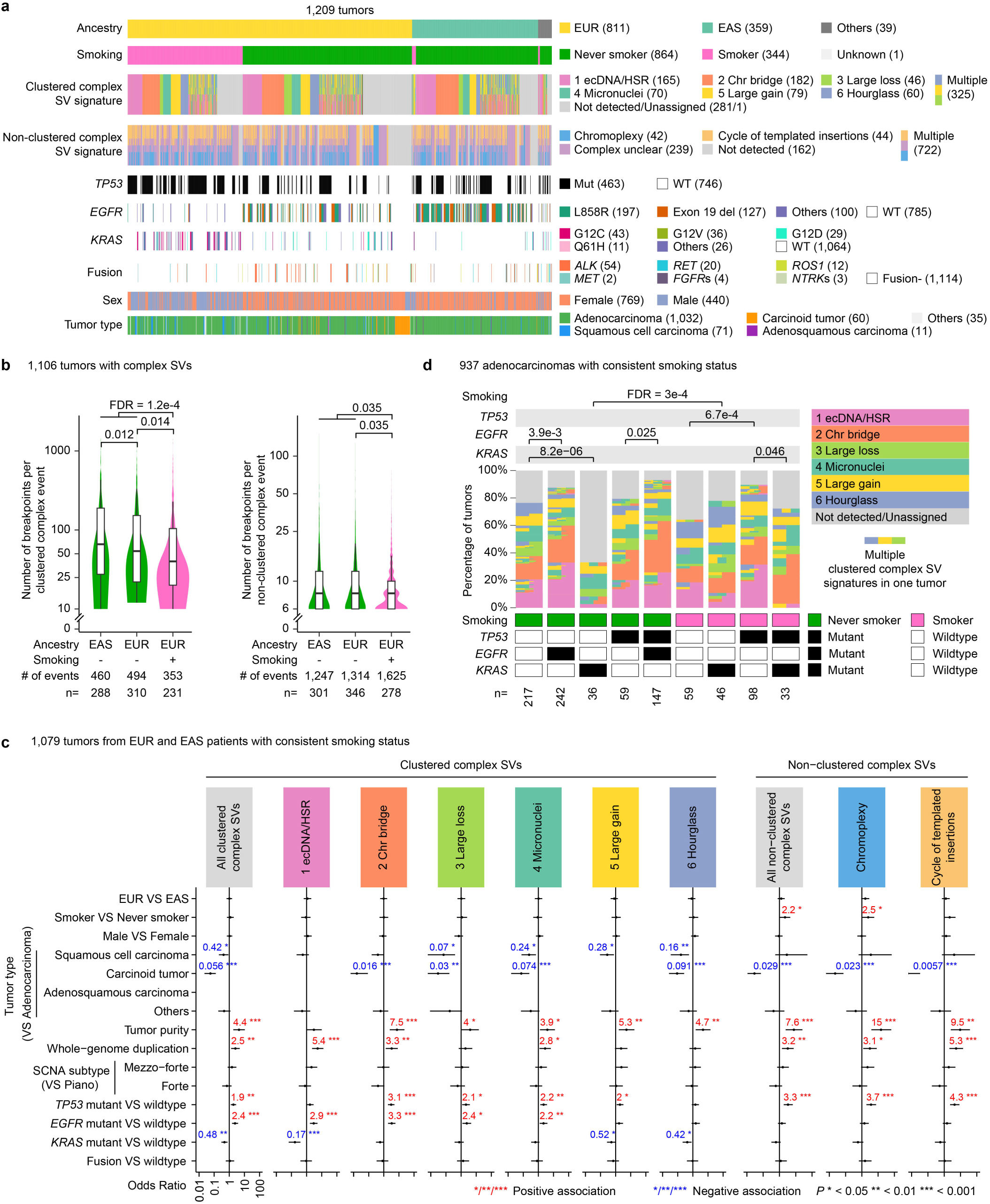
Clustered and non-clustered complex SVs. **a**, Overview of complex SVs in 1,209 tumors. The thin bars along the x-axis of each track represent individual tumors. For the 3^rd^ and 4^th^ signature tracks, the numbers in parentheses indicate the number of tumors with a unique complex SV signature of that type, while the “Multiple” groups in clustered and non-clustered complex SVs represent tumors with multiple complex SVs of different signatures. Therefore, numbers in parentheses may be smaller than those reported in the text, which include all tumors carrying that signature regardless of whether they have other complex SV signatures. **b**, Comparison of the numbers of breakpoints per clustered (left) and non-clustered (right) complex SV events across ancestry and smoking status, where n indicates the number of tumors in each group. Box plots within violins indicate the interquartile ranges and medians. Statistical comparisons are performed using two-sided Wilcoxon rank-sum tests with FDR correction; only FDRs < 0.05 are shown. **c**, Factors associated with clustered and non-clustered complex SV signatures using logistic regression. Odds ratios (ORs) are presented for significant factors in corresponding signatures. Error bars represent 95% confidence intervals. SCNA subtypes, piano (few SCNAs), mezzo-forte (enriched with arm-level amplification) and forte (dominated by whole-genome duplication), are defined according to our previous study^20^. **d**, Distributions of clustered complex SV signatures across tumors in selected groups. Each vertical column in the middle represents a tumor group defined by smoking status and mutation status of *TP53*, *EGFR* and *KRAS* in the bottom blocks. Each horizontal bar within a column in the middle represents one tumor. The height of each tumor may vary depending on the number of tumors (n) of the group. Tumors are color-coded based on clustered complex SV signatures with tumors exhibiting multiple SV signatures shown as horizontally segmented bars with different colors. Pairwise comparisons are performed for all clustered complex SVs combined between groups differing by only one factor using Fisher’s exact test with FDR correction; only FDRs < 0.05 are shown.

Many clinical and genetic factors differ between lung cancer types. For example, LCSS are mainly in males, whereas LCINS in females; carcinoid tumors are almost all from patients who have never smoked (**Fig. 1a**); *KRAS* and *TP53* mutations are enriched in LCSS, whereas *EGFR* mutations in LCSS^29^. To better assess factors that impact complex SVs, we performed a logistic regression using the presence or absence of complex SV signatures as the dependent variable and patient ancestry, sex, smoking status, tumor type, tumor purity, whole-genome duplication (WGD), SCNA subtype^20^, mutations of *TP53*, *EGFR*, *KRAS* and gene fusion status as independent variables. No strong multicollinearity was observed in the independent variables (**Extended Data** Fig. 2a and 2b). Tumor purity was positively associated with all complex SV signatures (**Fig. 2c**). Patient ancestry and sex had no effects on any complex SV signatures (**Fig. 2c**). Smoking was positively associated with chromoplexy (**Fig. 2c**). Carcinoid tumors had relatively quiet genomes with very few complex SVs (**Fig. 1b** and **2c**). WGD was positively associated with some clustered complex SVs and all non-clustered complex SVs (**Fig. 2c**). *TP53* mutations were associated with most of the complex SV signatures (**Fig. 2c**) as expected^40,43^. Intriguingly, *EGFR* mutations were positively associated with almost all clustered complex SV signatures, whereas *KRAS* mutations were negatively associated with ecDNA/HSR, large gain and hourglass signatures (**Fig. 2c**). Similar associations were found if only analyzing 602 high-clonality tumors, taking tumor purity into account (Methods) (**Extended Data** Fig. 2c). When LCSS and LCINS were considered separately, mutant *TP53* was positively associated with various types of complex SVs in both groups (**Extended Data** Fig. 3). *EGFR* mutations were positively associated with nearly all complex SV signatures in LCINS but not in LCSS, while *KRAS* mutations were negatively associated with clustered complex SVs in LCINS (**Extended Data** Fig. 3). We further stratified lung adenocarcinomas (LUADs) into distinct groups based on smoking status and mutation status of *TP53*, *EGFR* and *KRAS* (**Fig. 2d**). Among 217 and 242 LCINS (the left two groups in **Fig. 2d**) with wildtype *TP53* and *KRAS* but different *EGFR* status, clustered complex SVs were significantly more abundant in *EGFR*-mutant tumors (88.0%) than wildtype ones (76.5%). In LCINS with wildtype *TP53* and *EGFR*, the ones with *KRAS* mutation, had the lowest number of complex SVs (33.3%, **Fig. 2d**). Similarly, in *TP53*-mutant LCSS, those with also *KRAS*-mutations were associated with less clustered complex SVs than in those with no *KRAS* (72.7% and 89.8%, respectively).

In summary, LCSS had more complex SVs; however, they were more complex (more SV breakpoints per event) in LCINS. *EGFR* mutations were positively associated with complex SVs, and this effect was independent from other factors. The effect of *KRAS* mutations was the opposite of that observed for *TP53* and *EGFR* mutations.

### Simple SVs

After excluding complex SVs, 65,001 simple SVs (53.8 per tumor on average) remained in our cohort (**Supplementary Table S3**). We performed *de novo* simple SV signature decomposition using SigProfilerExtractor^44^, a non-negative-matrix-factorization-based *de novo* signature inference algorithm, rather than fitting SVs to COSMIC SV signatures. This is because COSMIC SV signatures do not separate complex and simple SVs, and four out of ten COSMIC SV signatures involve clustered SVs^45^. Furthermore, we classified simple SVs into 49 categories based on SV type and size to capture the effect of SV size better than in the COSMIC SV signatures^46^. For example, deletions were classified into 18 size categories, whereas COSMIC used 5 size categories. As a result, eight simple SV signatures were extracted using our approach (**Fig. 3a**). There were three deletion signatures (namely “Del1”, “Del2” and “Del3”), while COSMIC can only capture two (SV5 and SV7). Moreover, we found two tandem duplication (TD) signatures (namely “TD1” and “TD2”), as well as three other signatures including foldback inversions (“Fb inv”), intra-chromosomal translocations (“Intra tra”) and inter-chromosomal translocations (“Inter tra”) (**Fig. 3a**). This suggested that there were likely eight distinct mutational processes generating simple SVs in lung cancer. Our TD1, TD2 and Fb inv signatures corresponded to COSMIC SV3, SV1 and SV8 signatures, respectively. Our Intra tra and Inter tra signatures together resembled COSMIC SV2 signature. In our cohort, the most common simple SV signature was Del2 (16.6%), followed by Intra tra (16.0%) and Del3 (14.3%). The least abundant simple SV signature was TD1 (8.4%).

**Figure 3.**
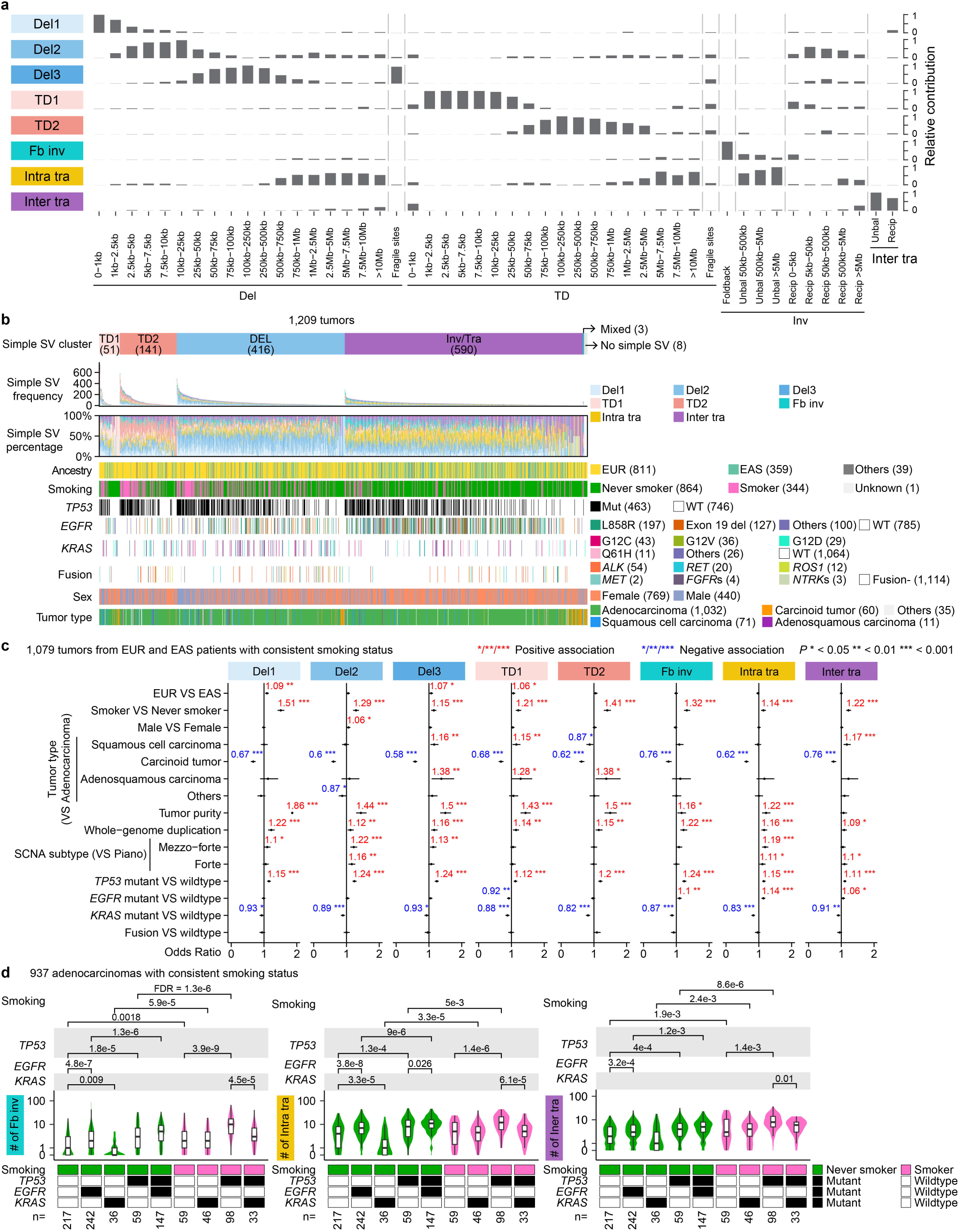
Simple SV signatures. **a**, Eight simple SV signatures detected in 1,209 tumors. The 49 SV categories used for signature extraction are shown on the x-axis. The eight simple SV signatures are shown on the left side of the y-axis. The height of each bar represents the proportion of SVs assigned to the corresponding signature. **b**, Five clusters of tumors based on the simple SV signatures (excluding tumors lacking simple SVs). Vertical lines represent individual tumors. **c**, Factors associated with simple SV signatures. ORs are presented for significant factors. Error bars represent 95% confidence intervals. **d**, Comparisons of simple SV signatures across selected groups. The violin plots illustrate the distributions of numbers of simple SVs for “Fb inv” signature on the left, “Intra tra” signature in the middle, and “Inter tra” signature on the right. Box plots within violins indicate the interquartile ranges and medians. Tumor group is defined by smoking status and mutation status of *TP53*, *EGFR* and *KRAS* in the bottom blocks. Statistical comparisons are performed between groups differing by only one factor using two-sided Wilcoxon rank-sum tests with FDR correction; only FDRs < 0.05 are shown.

We then performed unsupervised clustering on tumors based on the activities of simple SV signatures and found four major clusters and a minor cluster (**Fig. 3b**). Three of the four major clusters were enriched in TD1, TD2 and Del1,2,3 signatures respectively (**Fig. 3b**). The fourth major “Inv/Tra” cluster was the largest cluster composed of 590 (48.8%) tumors and was enriched in Fb inv, Intra tra and Inter tra signatures (**Fig. 3b**). The minor “Mixed” cluster, composed of three tumors, harbored different signatures and had no dominant signature. LCINS overall (*P*=2.1 × 10^-18^) and *EGFR*-mutated tumors (across all patients, *P*=7.4 × 10^-25^) were enriched in the Inv/Tra cluster (**Fig. 3b**). To investigate the effects of various clinical and genetic factors, we conducted linear regression analyses for the eight simple SV signatures. Tumor purity, smoking status and WGD were positively associated with all simple SV signatures; patient ancestry with Del1, Del3 and TD1; and sex with Del2 only (**Fig. 3c**). Carcinoid tumors had very stable genomes with fewer simple SVs (**Fig. 3c**). As expected, *TP53* mutations were positively associated with all signatures (**Fig. 3c**). Intriguingly, *EGFR* mutations were positively associated with inversions, intra-chromosomal and inter-chromosomal translocations (Fb inv, Intra tra and Inter tra), but negatively associated with TD1 (**Fig. 3c**). *KRAS* mutations were negatively associated with all signatures (**Fig. 3c**). Similar associations were found when we restricted the analysis to 606 high-clonality tumors from EUR and EAS patients with consistent smoking status (**Extended Data Fig. 4a**). When tumors were stratified by smoking status and mutation status of *TP53*, *EGFR* and *KRAS*, the above relationships largely held (**Extended Data** Fig. 4b and **Fig. 3d**). Interestingly, LCINS with SBS4 mutations had fewer simple SVs as well as fewer Del1, Del2, TD1, TD2 and Fb inv than LCSS with SBS4 mutations; whereas LCSS without SBS4 mutations had more simple SVs as well as Del1, Del3 and TD1 than LCINS without SBS4 mutations (**Extended Data Fig. 5**).

In summary, smoking, WGD and *TP53* mutations were positively associated with simple SVs. *EGFR* mutations were positively associated with foldback inversions, inter- and intra-chromosomal translocations. *KRAS* mutations were negatively associated with all simple SVs.

### SV breakpoint microhomology

SVs form through erroneous DNA damage repair and aberrant DNA replication^47^. The sequence features at SV breakpoints can allow us to infer SV forming mechanisms. Most breakpoints had 0 to 3 bp microhomology (**Extended Data Fig. 6**). Although both non-homologous end-joining (NHEJ) and alternative end-joining (alt-EJ) primarily produce SV breakpoints with microhomology within this range, with tens of thousands of SVs detected in our cohort, there were clear distinctions in microhomology patterns across different SV signatures. Overall, complex SVs often had the most abundant 1 bp microhomology suggesting NHEJ, whereas most simple SVs had dominant 2 bp microhomology suggesting alt-EJ (**Extended Data Fig. 6**). Specifically, the breakpoints in the two translocation signatures (Intra tra and Inter tra) were most abundant in 0 and 1 bp homology suggesting that NHEJ is the primary mechanism (**Extended Data Fig. 6**). Two bp microhomology dominated breakpoints in deletion, tandem duplication and inversion signatures (Del2, Del3, TD1, TD2 and Fb inv) (**Extended Data Fig. 6**) suggesting alt-EJ is more prevalent. Del1 was the only signature with the most frequent breakpoints being blunt ends (0 bp microhomology) (**Extended Data Fig. 6**). Microhomology of 2 bp was also abundant in Del1. These suggested that both NHEJ and alt-EJ contribute to Del1 formation. Noticeable portions of breakpoints in ecDNA/HSR, large gain, cycle of templated insertions and TD2 signatures had insertions of more than 10 bp (**Extended Data** Fig. 6), suggesting possible involvement of microhomology mediated break induced repair (MMBIR). Both MMBIR and alt-EJ might be relevant in the cycle of templated insertions (**Extended Data** Fig. 6).

### SV hotspots

The distribution of SV breakpoints across the genome is determined by a combined effect of DNA damage, repair and selection. SV breakpoint hotspots often represent regions with excessive DNA damage or SVs under positive selection. We split the genome into 1 Mb windows and calculated the percentage of tumors with SV breakpoints from individual SV signatures in each window. Different SV signatures displayed distinct breakpoint distributions (**Fig. 4**). Complex SVs were different from simple SVs. The hotspots in complex SVs primarily involved known oncogenes, whereas the hotspots in simple SVs were mostly fragile sites (i.e., genomic loci that are prone to DNA breaks^48^) as well as several oncogenes and tumor suppressors (**Fig. 4**). *MYC* and *MDM2* are among the most frequently amplified oncogenes in cancer including lung cancer^49^ and were frequently rearranged by complex SVs including ecDNA/HSR, chromatin bridge, large gain and hourglass signatures (**Fig. 4**). Out of 54 *EML4*-*ALK* fusions, 32 (59.3%) were produced by complex SVs including micronuclei, hourglass, chromoplexy and cycle of templated insertions; 14 (25.9%) and 8 (14.8%) were formed through unbalanced inversions (classified as Intra tra) and reciprocal inversions (classified as Del2, **Fig 3a**), respectively (**Supplementary Table S2**). As the most commonly deleted tumor suppressor^49^, *CDKN2A* was disrupted by many complex and simple SV signatures including large loss, chromoplexy, Del2, Del3 and Intra tra (**Fig. 4**). These together suggest functional convergence on a handful of major cancer driver genes from distinct molecular mechanisms of SV formation. The SV breakpoint distributions of ecDNA/HSR, chromatin bridge and large gain signatures were similar (**Fig. 4**) since these signatures often result in DNA copy gains^40^. Although large loss, micronuclei, hourglass and chromoplexy are signatures that commonly lead to loss of DNA, the SV breakpoint distributions were different (**Fig. 4**). For example, large loss had a hotspot at *MECOM*; micronuclei had a hotspot at *CCNE1*; hourglass had a hotspot at *MDM2*; whereas chromoplexy had none of these hotspots (**Fig. 4**). Intriguingly, the three deletion signatures had different SV breakpoint distributions. Large deletions (Del3) had many hotspots at fragile sites, but small (Del1) and median deletions (Del2) had none (**Fig. 4**). Hotspots of small TDs (TD1) did not overlap with any known oncogenes or tumor suppressors but were all fragile sites very similar to those in Del3 (**Fig. 4**) suggesting possible shared SV forming mechanisms of TD1 and Del3. There were very few hotspots in inter-chromosomal translocations (Inter tra), and the two major ones were due to transductions of L1 retrotransposons including a very active germline L1 element^30^ on chromosome 22 in the first intron of *TTC28* (**Fig. 4**). Interestingly, some SV hotspots were different between LCINS and LCSS. For example, *MDM2* was frequently rearranged by ecDNA/HSR in LCINS, but less frequent in LCSS (**Fig. 4**). *CDKN2A* was deleted by large loss and chromoplexy exclusively in LCINS, whereas *STK11* was frequently deleted by Del2 only in LCSS (**Fig. 4**).

**Figure 4.**
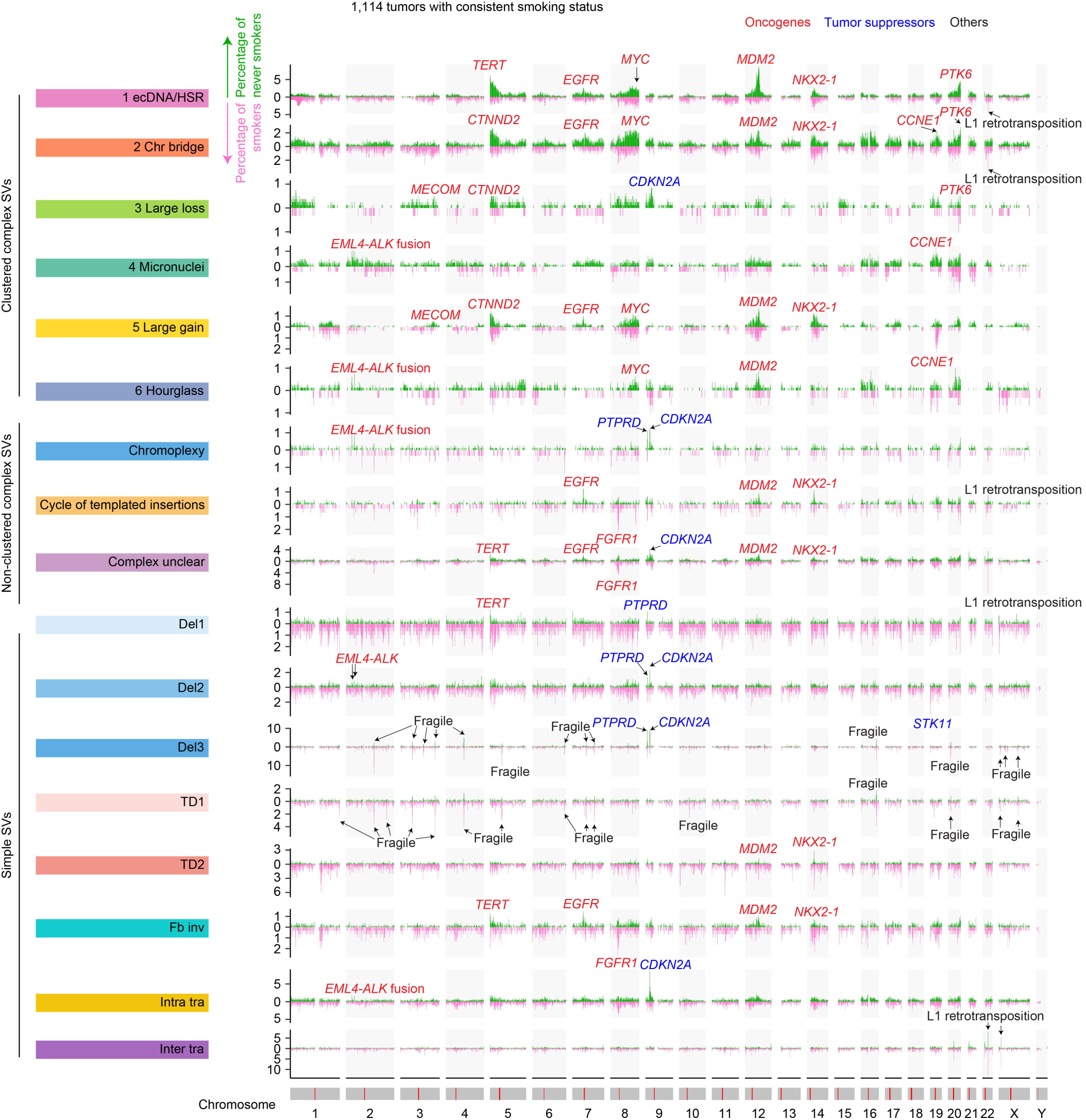
Genome-wide SV breakpoint distribution for each SV signature. Chromosomal structures are represented by grey bars at the bottom with red lines marking centromeres. Each vertical bar represents the percentage of tumors carrying SV breakpoints for a specific SV signature in the corresponding 1 Mb interval. Known oncogenes, tumor suppressors, fragile sites and L1 retrotransposition events are annotated in major peaks.

The breakpoint distribution across SV signatures could facilitate functional inferences of hotspot genes. *PTPRD* has been reported as a tumor suppressor^50^. However, it is a very large gene (2.3 Mb) and has been implicated as a fragile site similar to other large genes, such as *MACROD2*, *FHIT* and *WWOX*^22,51^. In our cohort, fragile site hotspots were shared between Del3 and TD1 but not present in any other signatures; *PTPRD* was not a hotspot in TD1 of which all hotspots were fragile sites; *PTPRD* was frequently rearranged in several SV signatures not involving fragile sites, such as chromoplexy, Del1 and Del2 (**Fig. 4**). These results combined indicate that *PTPRD* is likely a tumor suppressor instead of a fragile site. Interestingly, in some signatures, such as chromoplexy, Del2 and Del3, *PTPRD* was altered as frequently as *CDKN2A* (**Fig. 4**). However, it was the only frequently rearranged locus in Del1 in LCINS, whereas *CDKN2A* was the only major hotspot in intra-chromosomal translocations (Intra tra) in LCINS without frequent SVs at the *PTPRD* locus (**Fig. 4**).

In summary, SV breakpoints were not evenly distributed across the genome and differed across signatures. Distinct SV signatures shared several SV hotspots around well-known cancer driver genes, suggesting functional convergence. There were also noticeable differences between tumors from patients with different smoking statuses.

### Cancer-driving events

To further study SVs under positive selection, we comprehensively identified a total of 1,847 focal copy number changes of known cancer-driving genes as well as 2,284 driver mutations/indels in known driver genes in 1,114 tumors with consistent smoking status (**Fig. 5a** and **Supplementary Table 5**). Chromosome-arm level copy number changes were excluded so that the focal copy number changes of known driver genes should all result from SVs and represent cancer-driving SVs. *EGFR* mutations were enriched in LCINS and *KRAS* and *TP53* mutations in LCSS (**Fig. 5a**). Most driver genes were mutated more frequently in LCSS than LCINS (**Fig. 5a**). Some focal copy number changes, such as *CDKN2A* losses, *MDM2* gains and *EGFR* gains, were enriched in LCINS, whereas others, such as *STK11* losses, *NKX2-1* gains, *MCL1* gains and *FGFR1* gains, were enriched in LCSS (**Fig. 5a**). Furthermore, among LCINS, focal gains of *TERT*, *PTK6*, *EGFR*, *CTNND2* and *BRAF* as well as mutations in *TP53* and *RBM10* primarily occurred in tumors with *EGFR* mutations (**Extended Data Fig. 7**). Since *EGFR* mutation is the earliest genetic alteration in LCINS^52^ (ref companion manuscription on lung cancer evolution), these co-occurring genetic alterations suggest they may act as secondary oncogenic drivers specifically required for the oncogenic transformation of cells with *EGFR* mutations. We then performed linear regression analyses to model the number of driver events using patient ancestry, sex, smoking status, tumor histology, tumor purity, WGD status, SCNA subtype, mutation status of *TP53*, *EGFR* and *KRAS* as well as fusion status as independent variables. LCSS had more driver mutations (**Fig. 5b**). Although smoking was positively associated with SV burden, especially simple SVs (**Fig. 3c**), it was not associated with the number of driver SVs (**Fig. 5b**). Carcinoid tumors had fewer driver mutations and driver SVs (**Fig. 5b**). Interestingly, *TP53*, *EGFR* and *KRAS* mutations were all positively associated with overall driver mutations (**Fig. 5b**). In contrast, *EGFR*-mutant tumors had more driver SVs, whereas *KRAS*-mutant tumors had less driver SVs (**Fig. 5b**). Overall LCSS carried a higher number of driver mutations, a median of three, compared to one to two in LCINS (**Fig. 5c** left panel). Among LCINS, *EGFR*- and *KRAS*-mutant tumors had medians of two driver mutations, while the remaining tumors, including fusion-driven ones, had medians of one (**Fig. 5c** left panel). In contrast, when considering driver SVs, *EGFR*-mutant LCINS had a median of two such SVs, whereas all other groups had only one (**Fig. 5c** right panel). Among LCSS, *KRAS*-mutant tumors had the fewest driver SVs (median of one, **Fig. 5c** right panel). These results combined suggest that LCSS are mainly driven by mutations, whereas SVs play more important roles in tumorigenesis in *EGFR*-mutant LCINS. We further investigated the SV signatures in driver SVs. In LCINS, SV drivers were more likely to be complex events (**Fig. 5d** top left panel), such as ecDNA/HSR and chromatin bridge (**Fig. 5d** top right panel). In contrast, in LCSS, driver SVs were enriched in simple SVs (**Fig. 5d** bottom left panel), such as tandem duplications, inversions and translocations (**Fig. 5d** bottom right panel). Even though *STK11* focal losses preferentially occurred in LCSS (**Fig. 5a**), *STK11* in LCINS were more likely to be disrupted by complex SVs (**Fig. 5d** top left panel). In LCINS, no significant difference in SV signatures was observed between *EGFR*-mutant and wildtype tumors (**Extended Data Fig. 8**). These results collectively suggested that complex SVs play more important roles in tumorigenesis in LCINS.

**Figure 5.**
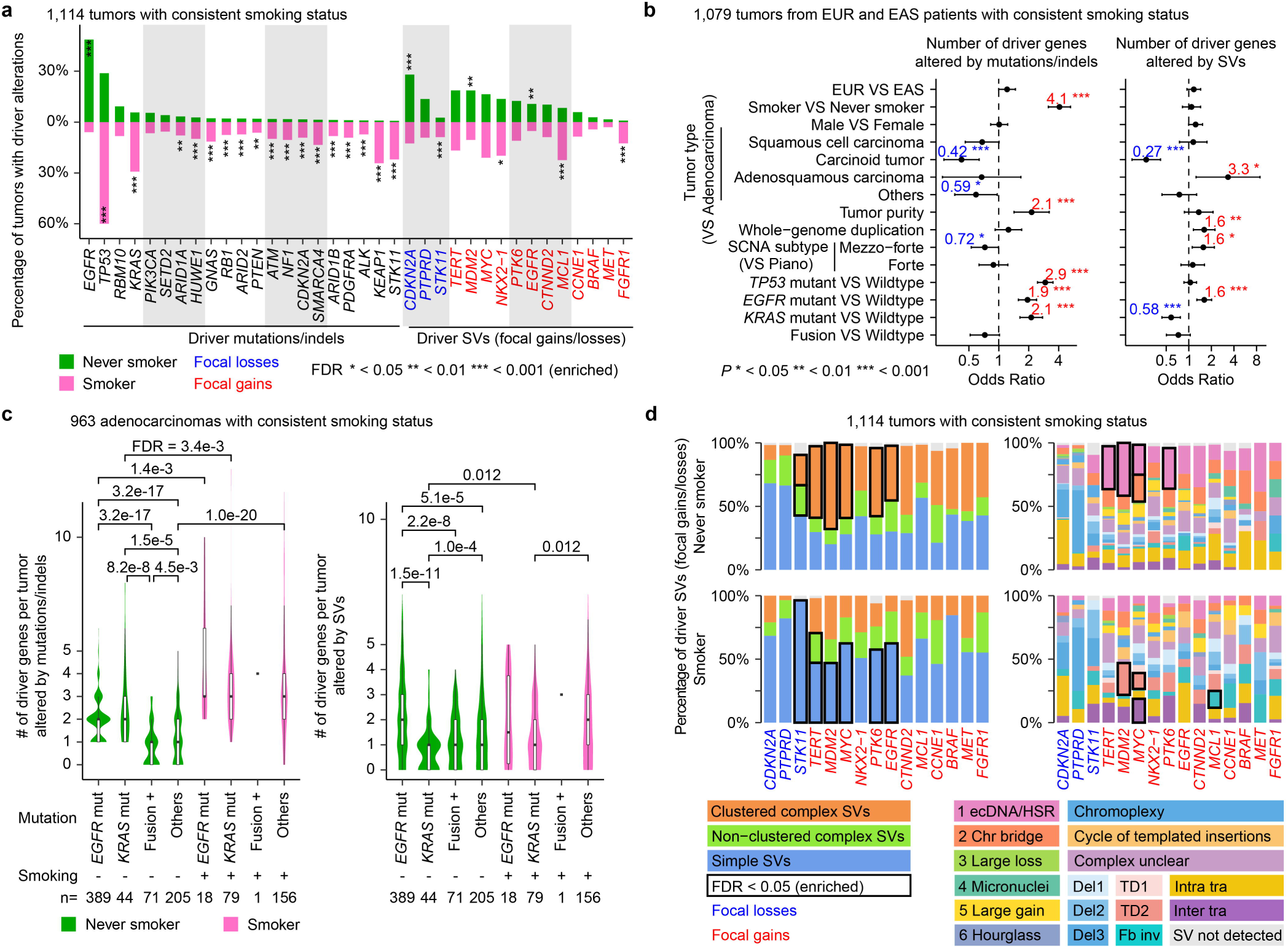
Cancer-driving mutations/indels and SVs. **a**, Proportions of tumors harboring mutations/indels and SVs in known cancer driver genes in LCINS and LCSS. “*”s that are in or above the green bars and in or below the pink bars represent alterations significantly enriched in LCINS and LCSS, respectively. Enrichment is assessed using Fisher’s exact test with FDR correction. **b**, Factors associated with the number of driver genes altered by mutations/indels (left) and by SVs (right). ORs are presented for significant factors. Error bars represent 95% confidence intervals. **c**, Numbers of driver genes altered by mutations/indels (left) and by SVs (right) across selected groups. Box plots within violins indicate the interquartile ranges and medians. Statistical comparisons are performed using two-sided Wilcoxon rank-sum tests with FDR correction; only FDRs < 0.05 are shown. **d**, SV signature composition of focal gains and losses in known cancer driver genes in LCINS (top) and LCSS (bottom). Colored bars represent SV types (left) and signatures (right). Enrichment of SV types or signatures comparing LCINS and LCSS is tested using Fisher’s exact test with FDR correction. Significantly enriched SV types and signatures are marked with black outlines.

As one of the commonly mutated genes in lung cancer, *EGFR* was mutated in 48.7% of the 809 LCINS. Among them, 86 (10.6%) had *EGFR* focal gains (driver SVs), which, in turn, were enriched with mutations/indels (80.2%, *P*=4.3 × 10^-10^, **Fig. 6a**). In contrast, 5.9% of LCSS had *EGFR* mutations/indels and a comparable fraction (6.3%, 1 out of 16) of tumors with focal gain of *EGFR* had mutations/indels (*P*=1, **Fig. 6a**). *EGFR* was frequently rearranged by several SV signatures, especially complex SVs (**Fig. 5d**), including ecDNA/HSR, chromatin bridge, large gain, cycle of templated insertions and Fb inv (**Fig. 4** and **Fig. 6b**). In addition, in tumors with *EGFR* mutations/indels and focal gains, the mutant allele was always amplified according to tumor purity and mutant allele fraction (**Fig. 6b**). Furthermore, *NKX2-1*, encoding a transcription factor to regulate cellular identity^53,54^ and recently recognized as an oncogene in LUAD^55^, was also frequently rearranged by multiple SV signatures such as ecDNA/HSR, chromatin bridge, large gain, cycle of templated insertions, TD2 and Fb inv (**Extended Data Fig. 9**).

**Figure 6.**
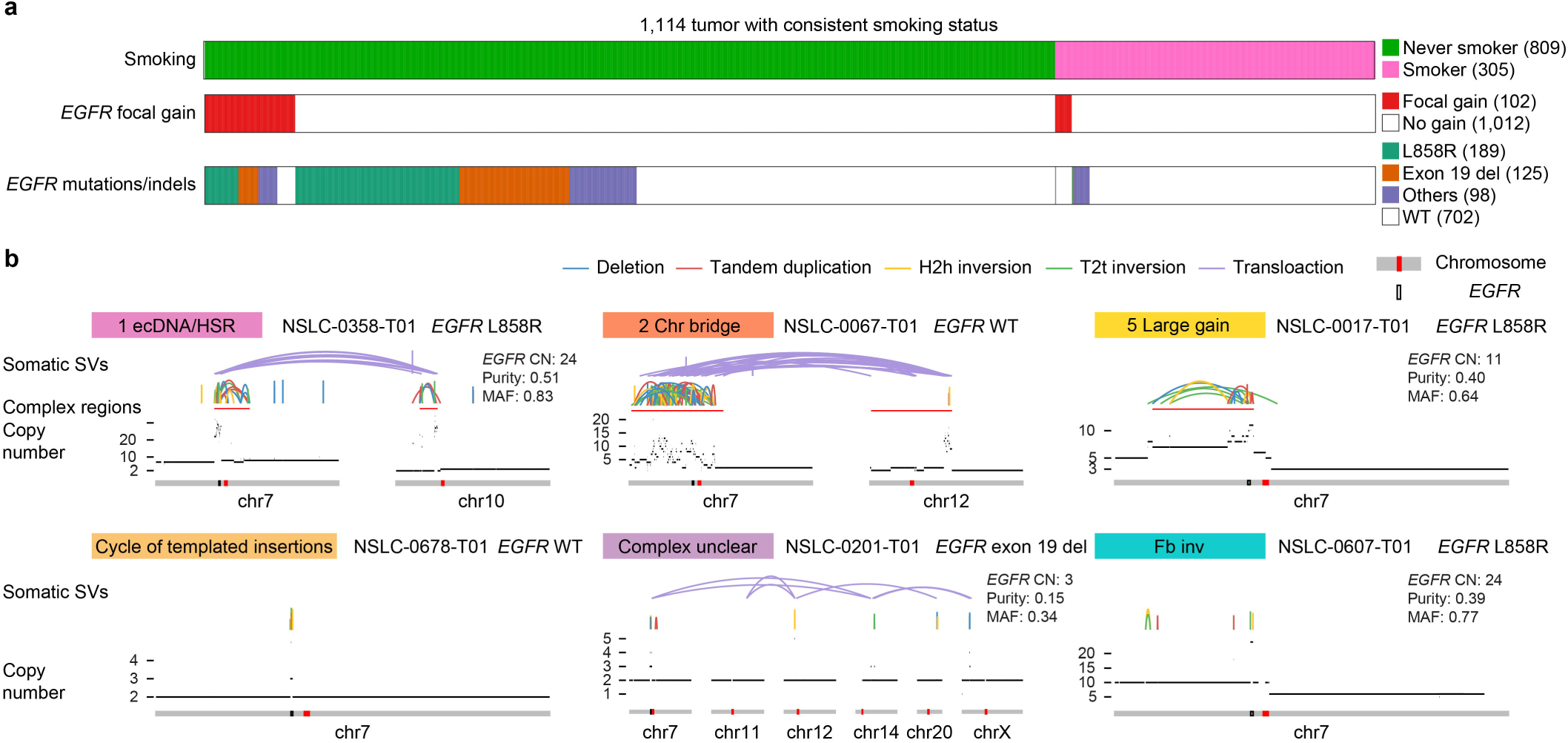
*EGFR*-associated SVs in LCINS and LCSS. **a**, Oncoprint plot showing *EGFR* focal gains and mutations/indels. **b**, Examples of SV signatures driving *EGFR* focal gains. Colored arcs represent SVs of different types. Red bars beneath the arcs mark the regions of clustered complex SVs. The copy number profiles are shown as black bars above the chromosome models. Centromere positions and *EGFR* genes are highlighted as red bars and black boxes within the grey chromosome ideograms. Sample IDs are displayed next to the corresponding SV signatures. *EGFR* copy number (CN), tumor purity, and mutant allele fraction (MAF) are listed for the cases with *EGFR* point mutations/indels.

In light of the findings within this study, we propose an updated oncogenic model for lung cancer (**Extended Data Fig. 10**). Most lung cancers carry two to five driver events (**Fig. 5c**) including point mutations, indels, fusions and DNA copy number changes. In LCSS epithelial cells, *KRAS* mutation is the tumor-initiating event caused by tobacco smoking. Secondary oncogenic alterations are primarily point mutations. One or two SVs per tumor serve as secondary drivers and are often simple SVs. In contrast, in LCINS cells, *EGFR* mutation arises through aging or endogenous processes as the first hit. One or two SVs, more frequently complex ones, are secondary drivers. In both LCSS and LCINS, malignant transformation occurs via specific combinations of genes being altered depending on the tumor-initiating events.

## Discussion

We performed a comprehensive study on somatic SVs in 1,209 lung cancers. We decomposed 16 signatures of complex and simple SVs. The distribution and microhomology of breakpoints revealed interesting insights into SV formation. Del3 and TD1 were associated with DNA breakage at fragile sites (**Fig. 4**). Complex SVs were more likely to form via NHEJ, whereas simple SVs were likely generated through alt-EJ (**Extended Data Fig. 6**).

We identified important factors associated with the SV panorama in lung cancer (**Fig. 2c** and **Fig. 3c**). They included smoking, WGD, and mutations in *TP53*, *EGFR* and *KRAS*. Patient ancestry and sex had little effect on genome instability. Intriguingly, although the kinase domain mutations in both *EGFR* and *KRAS* are well-known primary drivers of lung cancer, we found the opposite associations of these mutations with somatic SVs. Mutant *EGFR* was positively associated with complex SVs (**Fig. 2c**), inversions, intra- and inter-translocations (**Fig. 3c**), whereas mutant *KRAS* was not enriched with complex SVs, and was negatively associated with simple SVs (**Fig. 3c**). In a recent study, mutant *KRAS* was associated with more stable genomes in colorectal cancer^56^, which was consistent with our results in lung cancer. It is unlikely that the point mutations in *EGFR* and *KRAS* are the result of SVs, because *EGFR* and *KRAS* mutations are the earliest genetic alterations in lung cancer (ref companion manuscript on lung cancer evolution). Instead, it is more likely that *EGFR* and *KRAS* mutations shape the SV landscape, possibly involving different tumor cells of origin. The effects of these driver mutations can be through SV formation or selection. Indeed, *EGFR*-mutant tumors had more driver SVs, whereas *KRAS*-mutant tumors had less (**Fig 5b**). However, positive selection cannot entirely explain the SV patterns in *EGFR*-mutant tumors. Many known oncogenes, such as *MYC*, *MDM2* and *NKX2-1*, were amplified by various SV signatures which were positively associated with *EGFR* mutations, including ecDNA/HSR, chromatin bridge and large gain. *MDM2* and *NKX2-1* were also amplified by TD2 signature (**Fig. 4**). Circular ecDNA was studied in details in our companion paper^41^. If copy number gains of these genes are under positive selection in *EGFR*-mutant cells and SV burden is driven by positive selection, we would expect *EGFR* mutations to be positively associated with TD2 as well. However, *EGFR* mutations were not associated with TD2 in the full cohort (**Fig. 3c**) and negatively associated with TD2 in the high-clonality samples (**Extended Data Fig. 4a**). Therefore, the mechanisms of *EGFR* mutations shaping SV landscape may be due to a combined effect of *EGFR* mutations affecting SV formation and SVs affecting positive selection of altered cancer-driving genes. The detailed molecular mechanisms still require further experimental study.

Through detailed analysis of driver SVs and driver mutations, we revealed distinct patterns of driver events in LCSS and LCINS. The key findings were that LCSS had more driver mutations and LCINS had more SV drivers, which were more likely to be complex events (**Fig. 5c** and **5d**). There can be multiple possible reasons for these interesting patterns, including the positive selection of secondary drivers depending on the initial driver events or secondary drivers adapting to different microenvironments and immune modulation, which again warrant further study.

## Methods

### Ethics declarations

Since the National Cancer Institute only received de-identified samples and data from collaborating centers, had no direct contact or interaction with the study participants, and did not use or generate identifiable private information, Sherlock-*Lung* has been determined to constitute “Not Human Subject Research (NHSR)” based on the federal Common Rule (45 CFR 46; https://www.ecfr.gov/cgi-bin/ECFR?page=browse).

### Sample collection and processing

This study comprised 1,209 patients with histologically confirmed lung cancer primarily from Sherlock-*Lung* Project sourced from multiple institutions^29,30,34^ as well as publicly available datasets we previously collected^31–33,35–37^. Fresh-frozen tumor tissues were obtained alongside matched whole-blood samples or, when feasible, normal lung tissues sampled at least 3 cm from the tumor sites. All samples originated from treatment-naïve patients. Stringent inclusion criteria were uniformly applied^20^, ensuring minimum sequencing coverage of >40x for tumors and >25x for normal samples, limiting cross-sample contamination to <1% (evaluated by Conpair^57^), and maintaining sample relatedness below 0.2 (assessed by Somalier^58^). Samples exhibiting abnormal copy number profiles in matched normal tissues (determined by Battenberg^59^), samples with SBS7 (ultraviolet exposure-associated) or SBS31 (platinum chemotherapy-associated) mutational signatures, and those with fewer than 100 genomic alterations or fewer than 1,000 alterations combined with the low number of reads per clonal copy (NRPCC <10)^60^ were excluded. To avoid redundancy, only the highest-purity sample was selected in the case of multi-region sequencing. Furthermore, high-clonality tumor samples^30^ (n = 677) were defined as: (1) tumor purity > 0.3 to ensure adequate tumor cell content; (2) NRPCC ≥10 to enable reliable subclone detection; and (3) availability of high-confidence copy number profiles generated by Battenberg, excluding any samples that underwent refitting, thereby reducing potential artifacts from SCNAs. Genetic ancestry was assessed using verifyBamID^61^ and inferred through comparison with reference populations from the 1000 Genomes Project.

All coordinates in this study were based on the hg38 genome assembly. Gene annotation data were sourced from ENSEMBL (GRCh38.p13) (https://useast.ensembl.org/index.html).

### Variant calling and quality control

Genome-wide somatic alterations were identified using previously validated pipelines established in our Sherlock-*Lung* study^20,29–31^. Somatic mutations and small insertion-deletions (indels) were detected using a consensus approach involving four variant callers: Strelka (v2.9.10), MuTect, MuTect2, and TNscope (Sentieon Genomics v2023.08.03). Variants identified by at least three callers were retained. Germline contamination was excluded by filtering variants with allele frequency >0.001 in any of these germline databases (1000 Genomes Project, ExAC, Genome Aggregation Database). Functional annotations were assigned using ANNOVAR (v2020-06-08). Candidate driver genes were identified using the IntOGen pipeline19 (v2020.02.0123), incorporating seven complementary algorithms to highlight mutations under positive selective pressure. Allele-specific SCNAs were derived using the NGSpurity pipeline^62^, integrating Battenberg^59^ (v2.2.9), DPClust^59,63^ (v2.2.8), and ccube^64^ (v1.0). Joint estimation of tumor purity, ploidy, and clonal architecture was conducted^20,31^, followed by rigorous manual inspection and iterative refinement of purity and ploidy values.

Meerkat^65^ with suggested parameters and Manta^66^ with default parameters were used to call somatic SVs. To assess the sensitivity and specificity of SVs detection, thirty-five LUADs from the PCAWG study were used to benchmark SV calling strategies, since somatic SVs detected by PCAWG were robust with a 90% validation rate^17^. When comparing somatic SVs detected by PCAWG, Meerkat, and Manta, SVs were considered validated if the SV breakpoints were on the same chromosomes with the same orientations and within 50 bp. By the same token, SVs detected by Meerkat and Manta were merged if the SV breakpoints were on the same chromosomes with the same orientations and within 50 bp. Compared to individual algorithms, the union of SVs identified by the two algorithms showed the largest (2,884 somatic SVs) overlap with SVs detected by PCAWG (**Extended Data Fig. 11a**). In addition, there were 4,846 somatic SVs detected by Meerkat and Manta but not reported by PCAWG (**Extended Data Fig. 11a**). SCNA breakpoints were used to further evaluate the quality of SVs since unbalanced SVs should result in changes in DNA copy number. Three different distance cutoffs were used when comparing SV and SCNA breakpoints: 1 kb, 5 kb and 10 kb (**Extended Data Fig. 11b**). Among three groups of SVs, (a) only detected by Meerkat and Manta but not by PCAWG, (b) shared by Meerkat, Manta, and PCAWG and (c) only detected by PCAWG but not Meerkat or Manta, the fractions of SV breakpoints supported by SCNA breakpoints were comparable (**Extended Data Fig. 11b**). This suggests that the somatic SVs detected only by Meerkat and Manta but missed by PCAWG were as robust as those detected by PCAWG. Therefore, the union of somatic SVs detected by Meerkat and Manta was of high quality. Furthermore, some SV events were likely artifacts with limited support from SCNAs. To improve the reliability of SV calls in our dataset, we introduced additional filtering criteria tailored to our analysis: for Manta-specific SVs, at least 4 read pairs and 1 split read were required (**Extended Data Fig. 11c)**. For Meerkat-specific duplications that are less than 1,000 bp, additional >10 read pairs and >10 split reads were required (**Extended Data Fig. 11d)**. Microhomology length and insertion length around SV breakpoints were extracted from Meerkat and Manta.

SV breakpoints were annotated with protein-coding genes. SVs involving two distinct protein-coding genes with correct transcriptional orientations (5’-3’) were potential fusion genes. Fusions with 3’ partners being *ALK*, *RET*, *ROS1*, *MET, NTRKs* or with 5’ partners being *FGFRs* were considered driver fusions (**Supplementary Table S2**). To enhance reliability, only fusions with established partner genes^67^ were included in the analysis.

### SV signatures

Six clustered complex SV signatures were identified by Starfish^40^ (https://github.com/yanglab-computationalgenomics/Starfish) according to SV and SCNA patterns. The clustered complex SV signature cannot be extracted for one sample that has been annotated as “Unassigned” in the rest analysis as the SCNA data were not available. Non-clustered complex SVs were identified by ClusterSV^22^ (https://github.com/cancerit/ClusterSV) after removing clustered complex SVs.

After removing both clustered and non-clustered complex SVs, the remaining SVs were simple SVs. They were assigned into four primary types based on breakpoint locations and orientations: deletions, tandem duplications, inversions and translocations. Deletions and tandem duplications with breakpoints located in fragile regions were designated as fragile site deletions and fragile site tandem duplications, respectively. The remaining deletions and tandem duplications were further classified into 18 subgroups based on size. For inversions and translocations, they were further divided into reciprocal inversions, fold-back inversions, unbalanced inversions, reciprocal translocations and unbalanced translocations. Unbalanced inversions and reciprocal inversions were further split into 3 and 5 subgroups based on their sizes, respectively. Meanwhile, fold-back inversions, unbalanced translocations and reciprocal translocations stood as three distinct subcategories. In total, all simple SVs were classified into 49 subcategories. SigProfilerExtractor^44^ (https://github.com/AlexandrovLab/SigProfilerExtractor) with default parameters was employed to extract simple SV signatures.

Simple SVs were classified into their respective simple SV signatures as follows. Del1 signature included 0 – 2.5 kb deletions; Del2 signature included 2.5 kb – 25 kb deletions and all reciprocal inversions; Del3 signature included 25 kb – 750 kb deletions and fragile site deletions; TD1 signature included 0 – 50 kb tandem duplications and fragile site tandem duplications; TD2 signature included 50 kb – 5 Mb tandem duplications; Fb inv signature included foldback inversions; Intra tra signature included > 750 kb deletions, > 5 Mb tandem duplications and all unbalanced inversions; Inter tra signature included unbalanced inversions and reciprocal translocations. Tumors were clustered by the R package ConsensusClusterPlus^68^ based on the abundance of simple SV signatures. The clustering was performed with the following parameters: 80% samples resampling (pItem), 100% signatures resampling (pFeature), 50 resamplings (reps), agglomerative hierarchical clustering algorithm (clusterAlg) upon 1 – Pearson correlation distances (distance).

### Factors associated with SVs signatures and drivers

The list of cancer-driving genes was obtained from the Cancer Genome Atlas study^37^. Copy number alterations due to genome-level, chromosome level and chromosomal arm level alterations were discarded by considering the baseline copy numbers at genome, chromosome and chromosomal arm levels. The remainder of copy number alterations were focal events. Focal losses of tumor suppressors and focal gains of oncogenes were considered as SV drivers. Mutations and indels altering protein sequences were considered drivers. Logistic regression was used to identify associated factors for complex SV signatures, and linear regression was used to identify associated factors for simple SV signatures and the number of drivers. The presence and absence of complex SV signatures were used as dependent variables in logistic regression analyses. Similarly, the number of simple SVs and the number of drivers were used as dependent variables in linear regression. Tumors from African and South American patients were not analyzed because of their small sample size. All analyses were conducted in both the entire cohort and a high-clonality subset. The Fisher’s test assesses the differences in the distribution of tumors with and without clustered/non-clustered complex SVs across different groups; the two-sided Wilcoxon rank-sum test assesses the number of somatic SVs, simple SVs, breakpoints per complex event and signature-specific SVs in different groups; and the two-sided Student’s t-test evaluates the number of driver genes in different groups. Benjamini-Hochberg procedure was used for False Discovery Rate (FDR) correction. A cutoff of 0.05 was used for the significance level.

Clinical and genomic variables were encoded as numeric or binary values (e.g., smoking status, ancestry, WGD, driver mutations, fusion status), and a correlation matrix was generated for tumors from EUR and EAS patients with consistent smoking status. Pairwise Pearson correlations and Variance Inflation Factors (VIFs) were computed. Correlations > 0.8 or VIF > 5 were considered indicative of multicollinearity.

### Hotspots analysis

The reference genome was partitioned into non-overlapping bins of 1 Mb each. Within each bin, the number of tumors containing SV breakpoints was tallied for each SV signature. If a tumor carried multiple SV breakpoints of the same signature within the same bin, it was counted only once.

## Data availability

Normal and tumor-paired CRAM files for the WGS subjects of the Sherlock-*Lung* study and the EAGLE study have been deposited in dbGaP under the accession numbers phs001697.v2.p1 and phs002992.v1.p1, respectively. Detailed access information for the publicly available datasets can be found in **Supplementary Table 1**. Human reference genome GRCh38 was downloaded from the GATK resources at https://github.com/broadinstitute/gatk/blob/master/src/test/resources/large/Homo_sapiens_assembly38.fasta.gz.

## Code availability

All scripts used for data analysis and visualization are available at https://github.com/yanglab-computationalgenomics/Sherlock.SV.

## Acknowledgments

This work was supported by the Intramural Research Program of the National Institutes of Health (NIH) (project ZIACP101231 to MTL), the Anne Wojcicki Foundation (Grant Number: LC009 to MTL), the NIH grant R01CA269977 (LY), American Lung Association grant LCD-821295 (LY) and University of Chicago Medicine Comprehensive Cancer Center (LY). MD-G was awarded a fellowship within the “Generación D” initiative, Red.es, Ministerio para la Transformación Digital y de la Función Pública talent attraction (C005/24-ED CV1), funded by the European Union NextGenerationEU funds, through PRTR. The contributions of the NIH author(s) were made as part of their official duties as NIH federal employees, are in compliance with agency policy requirements, and are considered Works of the United States Government. However, the findings and conclusions presented in this paper are those of the author(s) and do not necessarily reflect the views of the NIH or the U.S. Department of Health and Human Services. We thank the study participants and the staff at Westat for their assistance with collecting samples and corresponding clinical data. This work used the computational resources of the NIH HPC Biowulf cluster (http://hpc.nih.gov).

## Author contributions

Conceptualization: TZ, LY, MTL; Methodology: YY, LY, BZ, LBA, DCW, MTL.; Formal analysis: YY, XZ, WZ, PHH, JSang, AK, JPM, CH, OWL, SS, KMJ, BZ, MD-G, TZ, LY, MAN, J.Shi; Pathology work: CL, MKB, WDT, LMS, PJ, RH, S-RY; Resources: CAH, MPW, KCL, ESE, JMS, MBS, SSY, MManczuk, JL, BS, AM, OS, DZ, IH, DM, MK, YB, BEGR, DCC, VG, PB, GL, PH, NR, DC, QL, SJC, VJ, SO, C-YC, ACP, MSavic, MSaha, CW, MTL; Data curation: PHH, TZ, MMiraftab, TVTT, OWL; Writing (original draft), YY, WP, LY, MTL; Writing (review and editing), YY, WP, LY, MTL; Visualization: YY, LY; Supervision: LY, MTL.

