## Extended Data Fig for "Panorama of Chromosomal Instability in Lung Cancer"

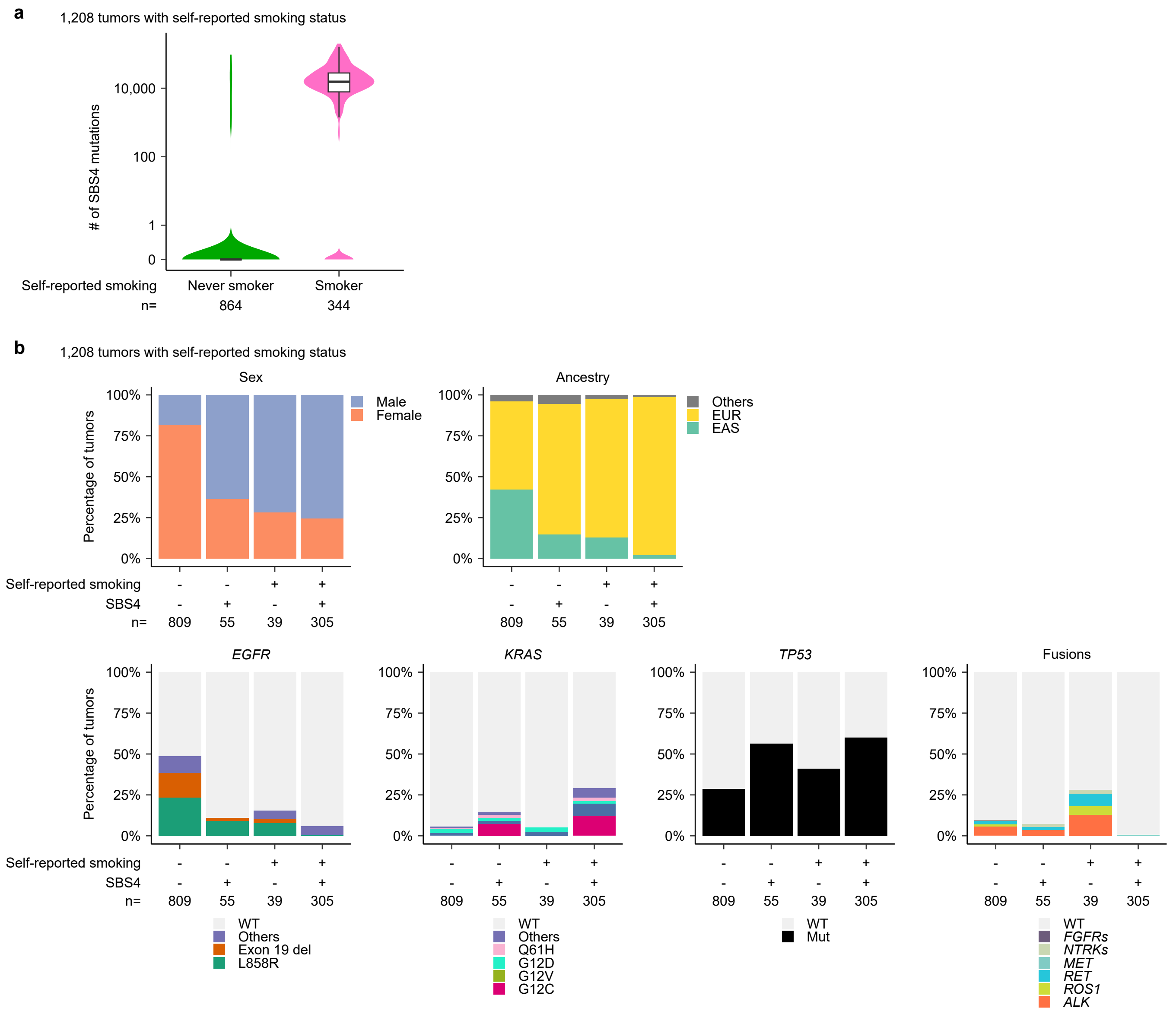
a
1,208 tumors with self-reported smoking status
### of SBS4 mutations
10,000
100
1
0
Self-reported smoking
Never smoker
Smoker
n=
864
344
b
1,208 tumors with self-reported smoking status
Sex
Percentage of tumors
100%
75%
50%
25%
0%
Male
Female
Ancestry
Others
EUR
EAS
Self-reported smoking - - + +
SBS4 - + - +
n= 809 55 39 305
EGFR
WT
Others
Exon 19 del
L858R
KRAS
WT
Others
Q61H
G12D
G12V
G12C
TP53
WT
Mut
Fusions
WT
FGFRs
NTRKs
MET
RET
ROS1
ALK

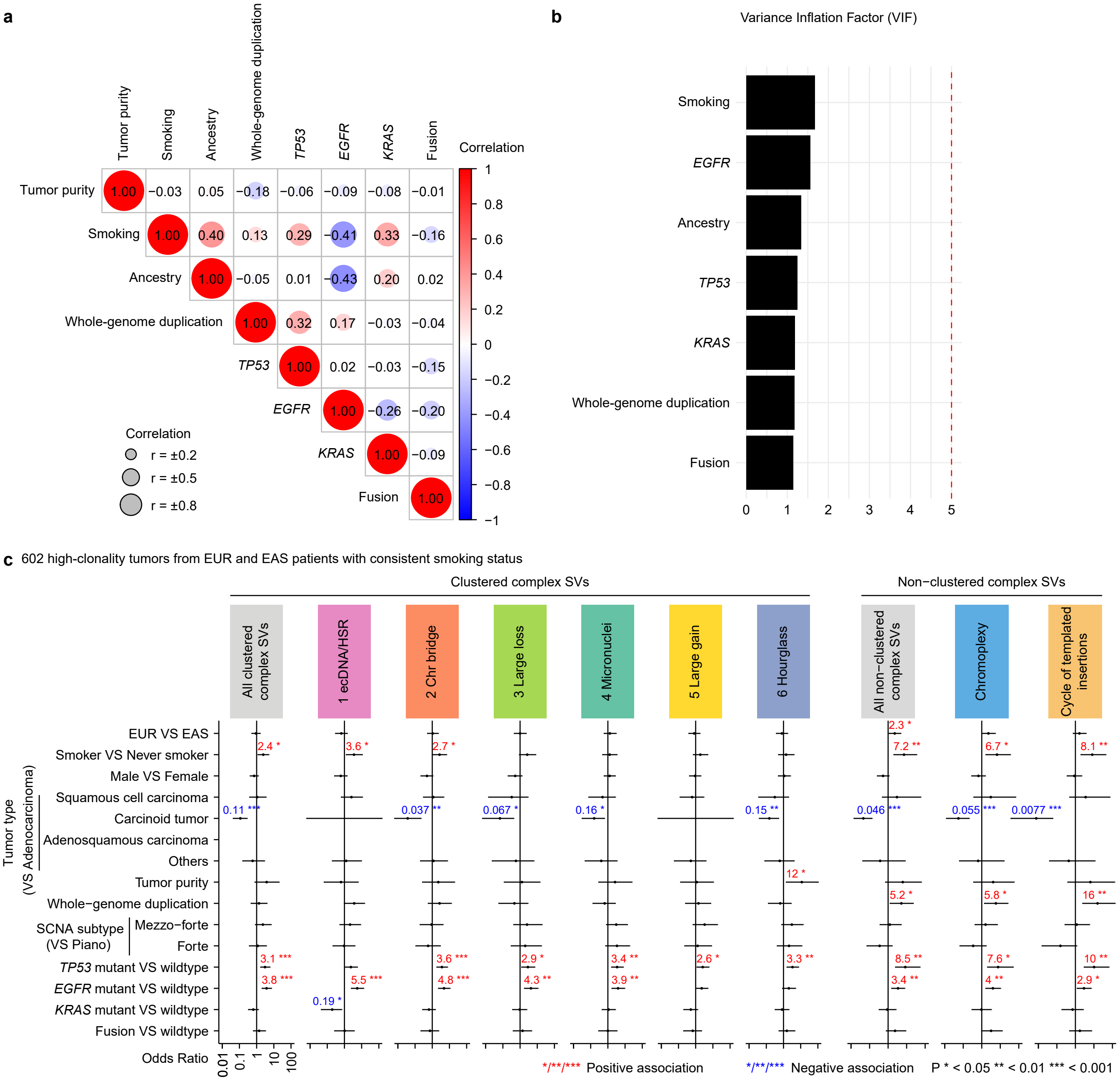
a
b
Variance Inflation Factor (VIF)
Correlation
c
602 high-clonality tumors from EUR and EAS patients with consistent smoking status
Clustered complex SVs
Non−clustered complex SVs
All clustered complex SVs
1 ecDNA/HSR
2 Chr bridge
3 Large loss
4 Micronuclei
5 Large gain
6 Hourglass
All non−clustered complex SVs
Chromoplexy
Cycle of templated insertions
Odds Ratio
*/**/*** Positive association
*/**/*** Negative association
P * < 0.05 ** < 0.01 *** < 0.001

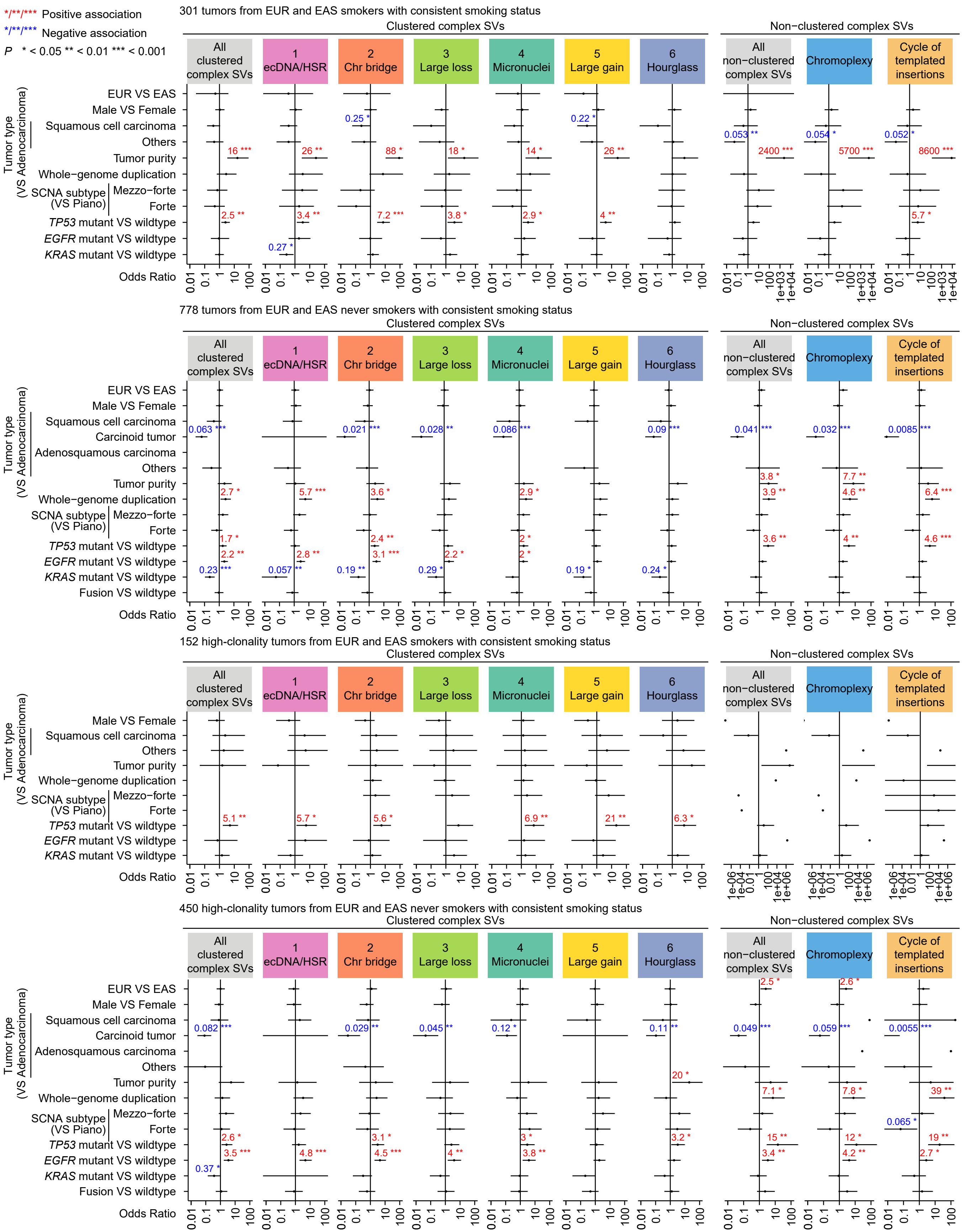

*/**/*** Positive association
*/**/*** Negative association
P * < 0.05 ** < 0.01 *** < 0.001
301 tumors from EUR and EAS smokers with consistent smoking status
Clustered complex SVs
Non−clustered complex SVs
All clustered complex SVs
1 ecDNA/HSR
2 Chr bridge
3 Large loss
4 Micronuclei
5 Large gain
6 Hourglass
All non−clustered complex SVs
Chromoplexy
Cycle of templated insertions
Tumor type (VS Adenocarcinoma)
EUR VS EAS
Male VS Female
Squamous cell carcinoma
Others
Tumor purity
Whole−genome duplication
SCNA subtype (VS Piano)
Mezzo−forte
Forte
TP53 mutant VS wildtype
EGFR mutant VS wildtype
KRAS mutant VS wildtype
Odds Ratio
16 ***
2.5 **
26 **
3.4 **
0.27 *
0.25 *
88 *
7.2 ***
18 *
3.8 *
14 *
2.9 *
0.22 *
26 **
4 **
0.053 **
2400 ***
0.054 *
5700 ***
0.052 *
8600 ***
5.7 *
0.01 0.1 1 10 100
0.01 0.1 1 10 100 1e+03 1e+04
778 tumors from EUR and EAS never smokers with consistent smoking status
Carcinoid tumor
Adenosquamous carcinoma
Fusion VS wildtype
0.063 ***
2.7 *
1.7 *
2.2 **
0.23 ***
5.7 ***
2.8 **
0.057 **
0.021 ***
3.6 *
2.4 **
3.1 ***
0.19 **
0.028 **
2.2 *
0.29 *
0.086 ***
2.9 *
2 *
2 *
0.19 *
0.09 ***
0.24 *
0.041 ***
3.8 *
3.9 **
3.6 **
0.032 ***
7.7 **
4.6 **
4 **
0.0085 ***
6.4 ***
4.6 ***
152 high-clonality tumors from EUR and EAS smokers with consistent smoking status
5.1 **
5.7 *
5.6 *
6.9 **
21 **
6.3 *
1e−06 1e−04 0.01 1 100 1e+04 1e+06
450 high-clonality tumors from EUR and EAS never smokers with consistent smoking status
0.082 ***
2.6 *
3.5 ***
0.37 *
4.8 ***
0.029 **
3.1 *
4.5 ***
0.045 **
4 **
0.12 *
3 *
3.8 **
0.11 **
20 *
3.2 *
2.5 *
0.049 ***
7.1 *
15 **
3.4 **
2.6 *
0.059 ***
7.8 *
12 *
4.2 **
0.0055 ***
39 **
0.065 *
19 **
2.7 *

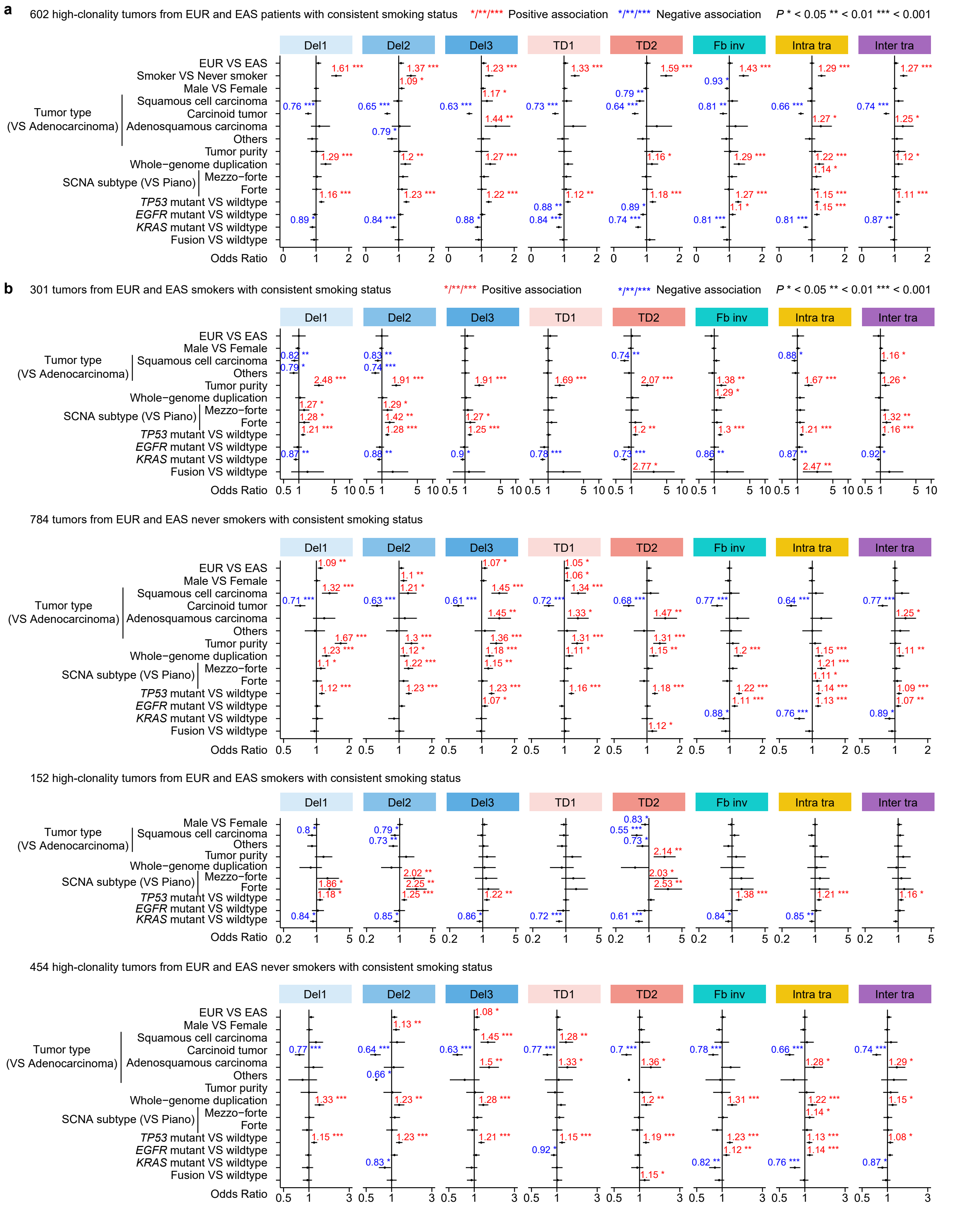
a
602 high-clonality tumors from EUR and EAS patients with consistent smoking status
*/**/*** Positive association
*/**/*** Negative association
P * < 0.05 ** < 0.01 *** < 0.001
Del1
Del2
Del3
TD1
TD2
Fb inv
Intra tra
Inter tra
EUR VS EAS
Smoker VS Never smoker
Male VS Female
Tumor type (VS Adenocarcinoma)
Squamous cell carcinoma
Carcinoid tumor
Adenosquamous carcinoma
Others
Tumor purity
Whole−genome duplication
SCNA subtype (VS Piano)
Mezzo−forte
Forte
TP53 mutant VS wildtype
EGFR mutant VS wildtype
KRAS mutant VS wildtype
Fusion VS wildtype
Odds Ratio
b
301 tumors from EUR and EAS smokers with consistent smoking status
784 tumors from EUR and EAS never smokers with consistent smoking status
152 high-clonality tumors from EUR and EAS smokers with consistent smoking status
454 high-clonality tumors from EUR and EAS never smokers with consistent smoking status

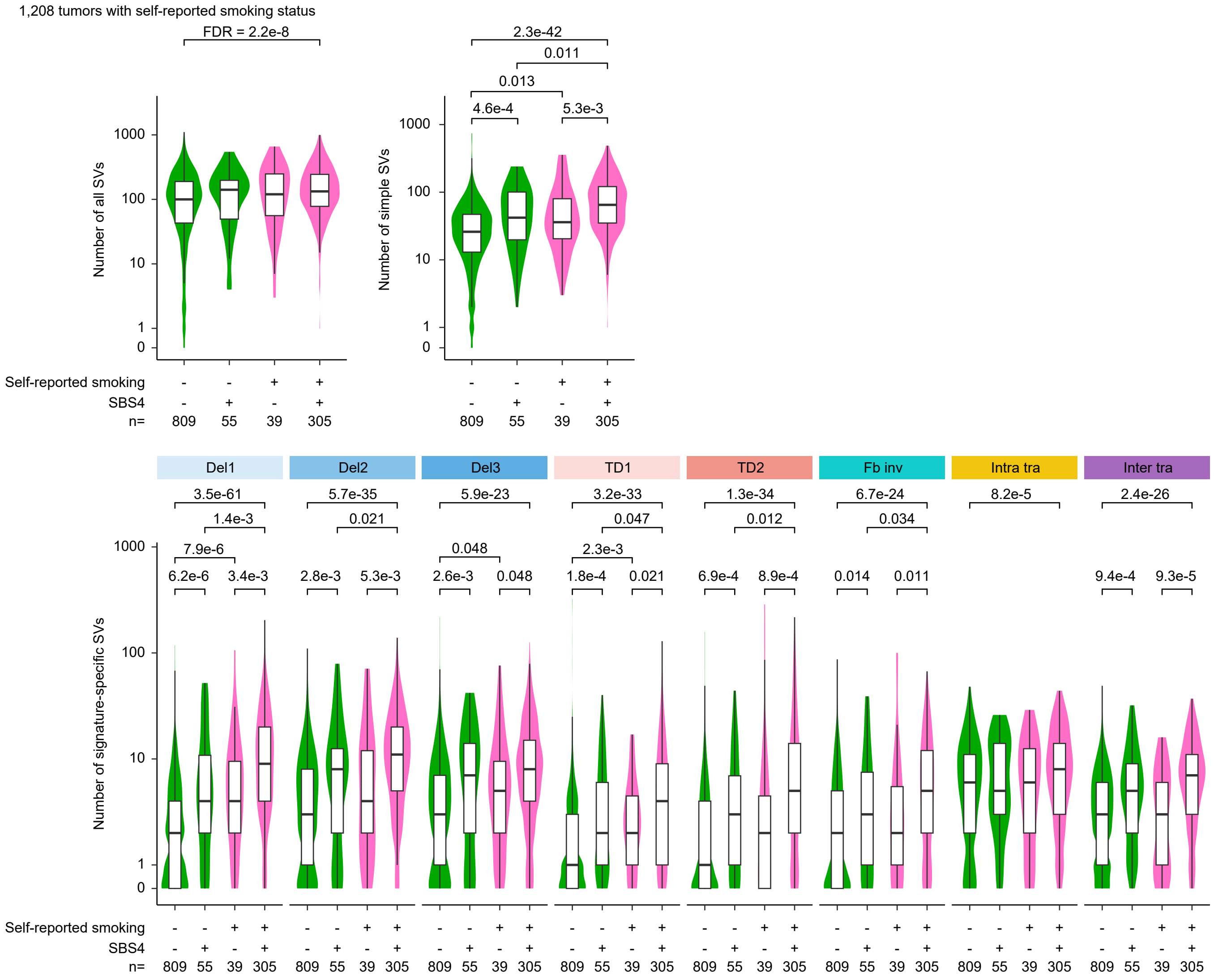

1,208 tumors with self-reported smoking status
FDR = 2.2e-8
Number of all SVs
2.3e-42
0.011
0.013
4.6e-4
5.3e-3
Number of simple SVs
Self-reported smoking
SBS4
n=
809
55
39
305
Del1
Del2
Del3
TD1
TD2
Fb inv
Intra tra
Inter tra
3.5e-61
1.4e-3
7.9e-6
6.2e-6
3.4e-3
5.7e-35
0.021
2.8e-3
5.3e-3
5.9e-23
0.048
2.6e-3
0.048
3.2e-33
0.047
2.3e-3
1.8e-4
0.021
1.3e-34
0.012
6.9e-4
8.9e-4
6.7e-24
0.034
0.014
0.011
8.2e-5
2.4e-26
9.4e-4
9.3e-5
Number of signature-specific SVs

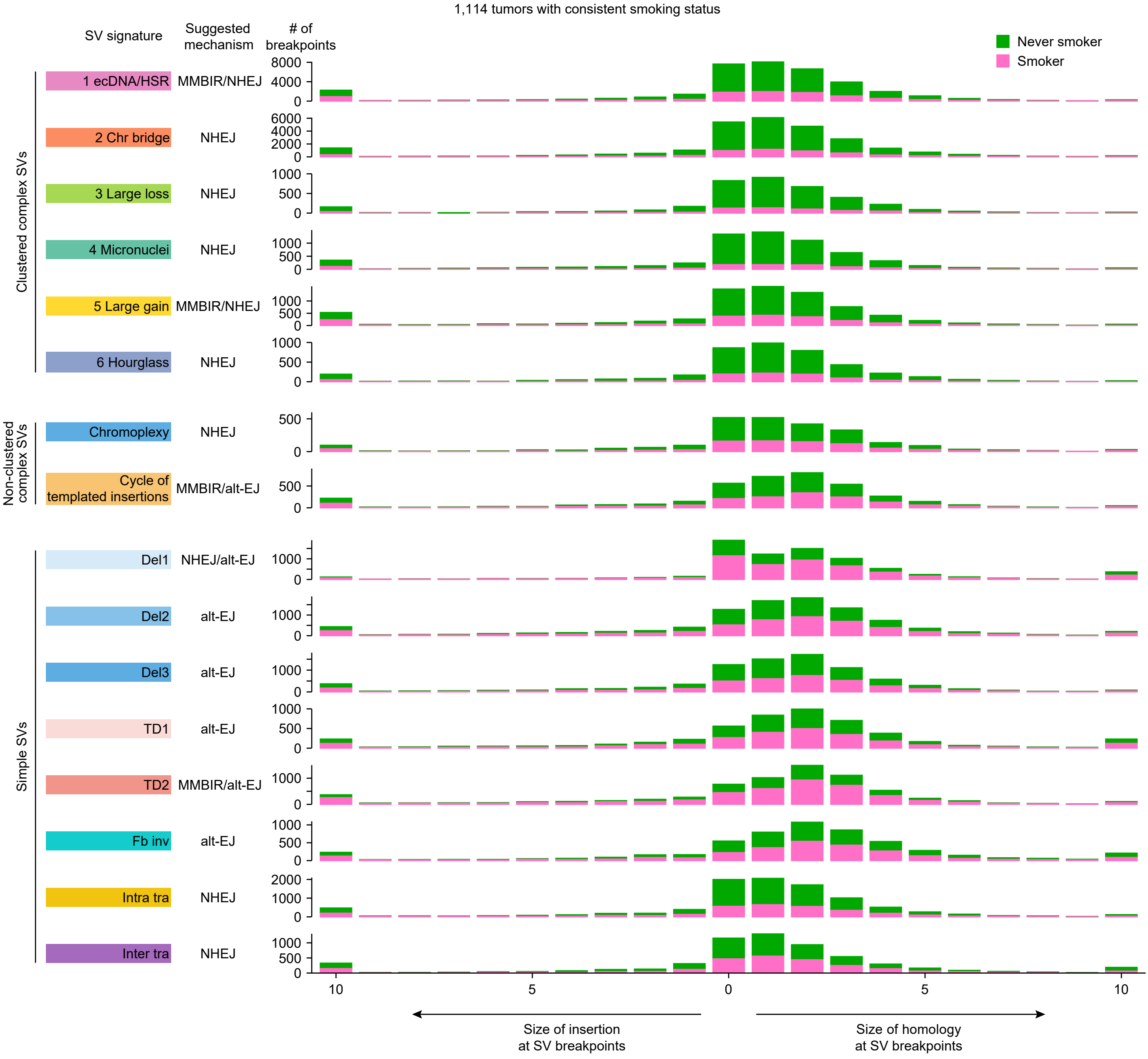

1,114 tumors with consistent smoking status
SV signature
Suggested mechanism
### of breakpoints
Never smoker
Smoker
Clustered complex SVs
1 ecDNA/HSR
MMBIR/NHEJ
2 Chr bridge
NHEJ
3 Large loss
NHEJ
4 Micronuclei
NHEJ
5 Large gain
MMBIR/NHEJ
6 Hourglass
NHEJ
Non-clustered complex SVs
Chromoplexy
NHEJ
Cycle of templated insertions
MMBIR/alt-EJ
Simple SVs
Del1
NHEJ/alt-EJ
Del2
alt-EJ
Del3
alt-EJ
TD1
alt-EJ
TD2
MMBIR/alt-EJ
Fb inv
alt-EJ
Intra tra
NHEJ
Inter tra
NHEJ
10
5
0
5
10
Size of insertion at SV breakpoints
Size of homology at SV breakpoints

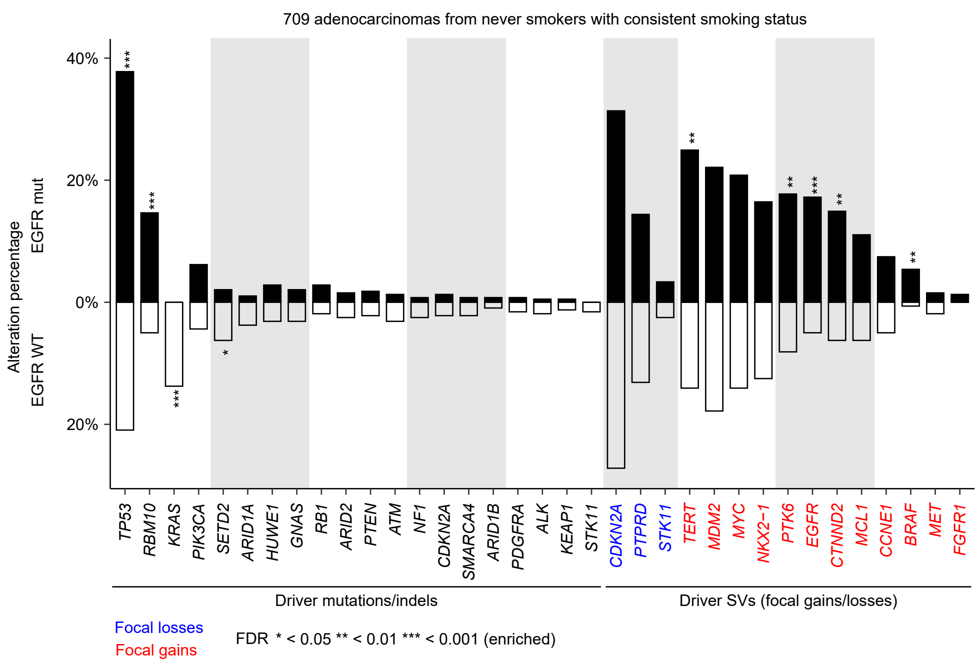
709 adenocarcinomas from never smokers with consistent smoking status
Alteration percentage
EGFR mut
EGFR WT
40%
20%
0%
20%
TP53
RBM10
KRAS
PIK3CA
SETD2
ARID1A
HUWE1
GNAS
RB1
ARID2
PTEN
ATM
NF1
CDKN2A
SMARCA4
ARID1B
PDGFRA
ALK
KEAP1
STK11
CDKN2A
PTPRD
STK11
TERT
MDM2
MYC
NKX2−1
PTK6
EGFR
CTNND2
MCL1
CCNE1
BRAF
MET
FGFR1
Driver mutations/indels
Driver SVs (focal gains/losses)
Focal losses
Focal gains
FDR * < 0.05 ** < 0.01 *** < 0.001 (enriched)

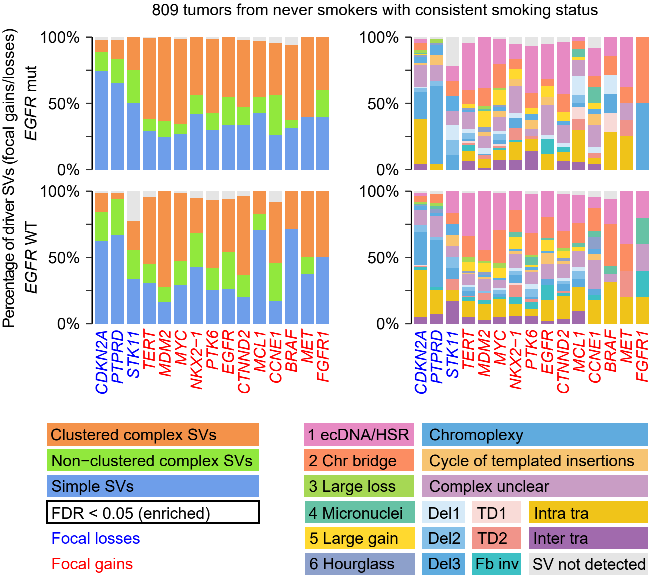
809 tumors from never smokers with consistent smoking status
Percentage of driver SVs (focal gains/losses)
EGFR mut
EGFR WT
100%
50%
0%
CDKN2A
PTPRD
STK11
TERT
MDM2
MYC
NKX2−1
PTK6
EGFR
CTNND2
MCL1
CCNE1
BRAF
MET
FGFR1
Clustered complex SVs
Non−clustered complex SVs
Simple SVs
FDR < 0.05 (enriched)
Focal losses
Focal gains
1 ecDNA/HSR
2 Chr bridge
3 Large loss
4 Micronuclei
5 Large gain
6 Hourglass
Chromoplexy
Cycle of templated insertions
Complex unclear
Del1
Del2
Del3
TD1
TD2
Fb inv
Intra tra
Inter tra
SV not detected

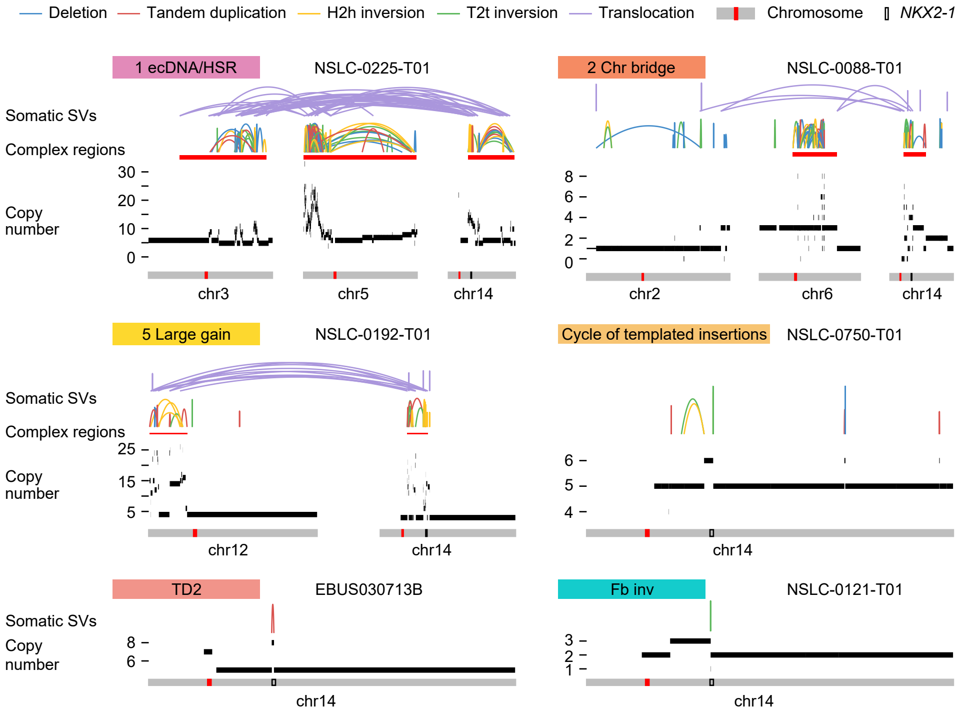

Deletion
Tandem duplication
H2h inversion
T2t inversion
Translocation
Chromosome
NKX2-1
1 ecDNA/HSR
NSLC-0225-T01
Somatic SVs
Complex regions
Copy number
30
20
10
0
chr3
chr5
chr14
2 Chr bridge
NSLC-0088-T01
8
6
4
2
0
chr2
chr6
chr14
5 Large gain
NSLC-0192-T01
Somatic SVs
Complex regions
Copy number
25
15
5
chr12
chr14
Cycle of templated insertions
NSLC-0750-T01
6
5
4
chr14
TD2
EBUS030713B
Somatic SVs
Copy number
8
6
chr14
Fb inv
NSLC-0121-T01
3
2
1
chr14

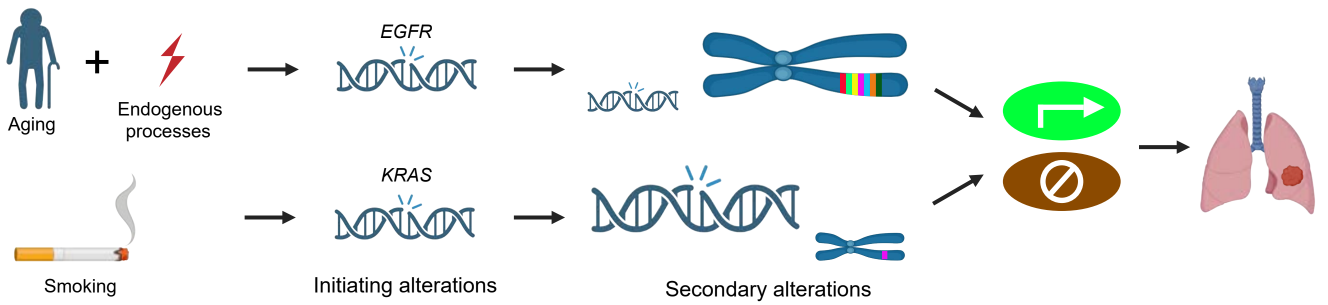
Aging
Endogenous processes
EGFR
KRAS
Smoking
Initiating alterations
Secondary alterations

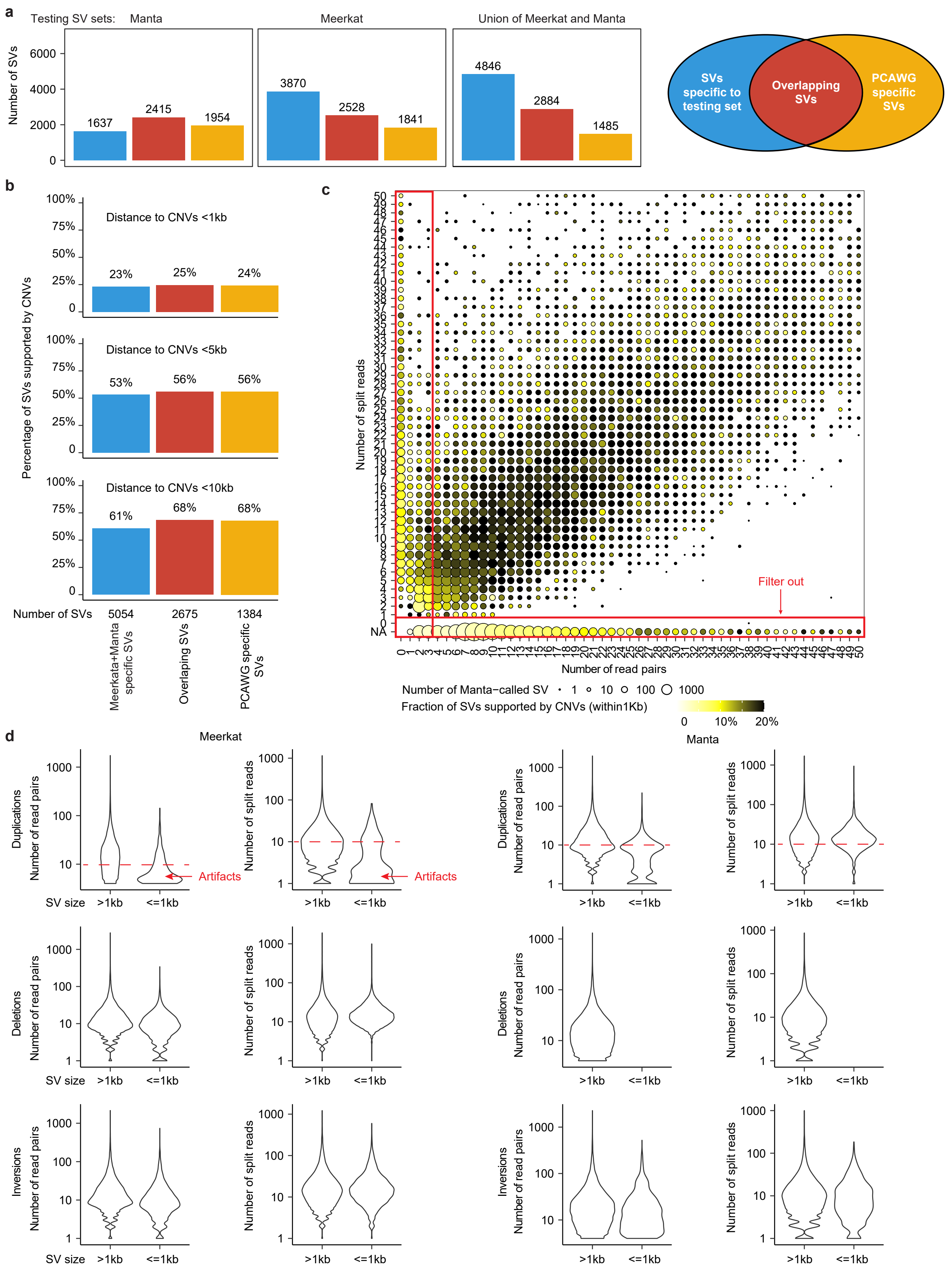
a
Testing SV sets: Manta
Meerkat
Union of Meerkat and Manta
Number of SVs
6000
4000
2000
0
1637
2415
1954
3870
2528
1841
4846
2884
1485
SVs specific to testing set
Overlapping SVs
PCAWG specific SVs
b
Percentage of SVs supported by CNVs
100%
75%
50%
25%
0
Distance to CNVs <1kb
23%
25%
24%
Distance to CNVs <5kb
53%
56%
56%
Distance to CNVs <10kb
61%
68%
68%
Number of SVs
5054
2675
1384
Meerkata+Manta specific SVs
Overlaping SVs
PCAWG specific SVs
c
Number of split reads
Number of read pairs
Filter out
NA
Number of Manta−called SV
1
10
100
1000
Fraction of SVs supported by CNVs (within1Kb)
0
10%
20%
d
Meerkat
Manta
Duplications
Deletions
Inversions
Number of read pairs
Number of split reads
1000
100
10
1
Artifacts
Artifacts
SV size
>1kb
<=1kb
